# Household secondary attack rates of SARS-CoV-2 by variant and vaccination status: an updated systematic review and meta-analysis

**DOI:** 10.1101/2022.01.09.22268984

**Authors:** Zachary J. Madewell, Yang Yang, Ira M. Longini, M. Elizabeth Halloran, Natalie E. Dean

**Author notes:** Correspondence to: Zachary J. Madewell, Department of Biostatistics, University of Florida, PO Box 117450, Gainesville, FL 32611.

## Abstract

We previously reported a household secondary attack rate (SAR) for SARS-CoV-2 of 18.9% through June 17, 2021. To examine how emerging variants and increased vaccination have affected transmission rates, we searched PubMed from June 18, 2021, through January 7, 2022. Meta-analyses used generalized linear mixed models to obtain SAR estimates and 95%CI, disaggregated by several covariates. SARs were used to estimate vaccine effectiveness based on the transmission probability for susceptibility (*VE*_*S,p*_), infectiousness (*VE*_*I,p*_), and total vaccine effectiveness (*VE*_*T,p*_). Household SAR for 27 studies with midpoints in 2021 was 35.8% (95%CI, 30.6%-41.3%), compared to 15.7% (95%CI, 13.3%-18.4%) for 62 studies with midpoints through April 2020. Household SARs were 38.0% (95%CI, 36.0%-40.0%), 30.8% (95%CI, 23.5%-39.3%), and 22.5% (95%CI, 18.6%-26.8%) for Alpha, Delta, and Beta, respectively. *VE*_*I,p*_, *VE*_*S,p*_, and *VE*_*T,p*_ were 56.6% (95%CI, 28.7%-73.6%), 70.3% (95%CI, 59.3%-78.4%), and 86.8% (95%CI, 76.7%-92.5%) for full vaccination, and 27.5% (95%CI, -6.4%-50.7%), 43.9% (95%CI, 21.8%-59.7%), and 59.9% (95%CI, 34.4%-75.5%) for partial vaccination, respectively. Household contacts exposed to Alpha or Delta are at increased risk of infection compared to the original wild-type strain. Vaccination reduced susceptibility to infection and transmission to others.

**Summary:** Household secondary attack rates (SARs) were higher for Alpha and Delta variants than previous estimates. SARs were higher to unvaccinated contacts than to partially or fully vaccinated contacts and were higher from unvaccinated index cases than from fully vaccinated index cases.

## Introduction

A previous systematic review and meta-analysis of household transmission of SARS-CoV-2 published through June 17, 2021 reported an overall secondary attack rate (SAR) of 18.9% (95% CI, 16.2%-22.0%) [1]. Emerging variants of concern and increased vaccination have affected transmission rates. Delta (B.1.617.2) became the predominant variant in many parts of the world and Omicron (B.1.1.529) poses additional challenges given its high level of spike mutations and increased potential for transmissibility [2, 3]. Other variants of concern include Alpha (B.1.1.7), Beta (B.1.351), and Gamma (P.1).

Vaccine efficacies against symptomatic disease and death have been demonstrated in randomized controlled trials [4] and vaccine effectiveness has been corroborated in large observational studies in Denmark [5], Israel [6], and the United Kingdom [7]. Household studies can supplement efficacy trials to determine vaccine effectiveness. Vaccine studies based on secondary attack rates (SARs) can be used to estimate the protective effectiveness of a vaccine in vaccinated susceptible contacts compared to unvaccinated susceptible contacts who are exposed to an infected index case (*VE*_*S,p*_) [8, 9]. Household studies also enable estimation of vaccine effectiveness in reducing infectiousness (*VE*_*I,p*_) by comparing SARs from vaccinated and from unvaccinated index cases to household contacts, which was done for pertussis [10]. Total vaccine effectiveness (*VE*_*T,p*_), or the combined effect of direct vaccine protection and indirect vaccine effectiveness, can also be estimated. It is unknown how effective the SARS-CoV-2 vaccines are in reducing susceptibility and infectiousness in the confines of the household where there is prolonged close contact between household members and index cases. Here, we aggregate evidence of household contact tracing studies to evaluate SARs for variants of concern and by index case or contact vaccination status.

## Methods

This study followed the Preferred Reporting Items for Systematic Reviews and Meta-analyses (PRISMA) reporting guideline using the same definitions and eligibility criteria as our original study [11]. Our last review identified studies published through June 17, 2018 [1]. Herein, we searched PubMed and reference lists of eligible studies between June 18, 2021, and January 7, 2022, with no restrictions on language, study design, or place of publication. Search terms were: “SARS-CoV-2”, “COVID-19”, “severe acute respiratory syndrome”, “SARS”, “SARS-CoV”, “coronavirus”, “variant”, “vaccination”, “immunization”, “secondary attack rate”, “secondary infection rate”, “household”, “family contacts”, “close contacts”, “index case”, “contact transmission”, “contact attack rate”, and “family transmission” (S1 Table). Pre-prints were included. Citations were managed in EndNote 20 (Thomson Reuters, Toronto, CA).

Articles with original data that reported at least 2 of the following factors were included: number of infected household contacts, total number of household contacts, and household secondary attack rates. Studies that reported infection prevalence, included populations that overlapped with another included study, and tested contacts using antibody tests only or using antibody tests and another test but did not disaggregate SARs by test were excluded. We first screened studies by titles and abstracts to identify potential studies for inclusion. That reviewer then evaluated full-text articles and selected those that met the inclusion criteria.

For this study, we extracted the following information: first author, location, index case identification period, number of index cases, index case symptom status, household/family contact type, test used to diagnose contacts, universal/symptomatic testing, number of tests per contact, and follow-up duration. We also extracted the number of infected household contacts and total number of household contacts and disaggregated by covariates including variant, index case vaccination status, household contact vaccination status, and vaccine type.

To examine temporal patterns, we assessed household SARs by index case identification period midpoint. We restricted this analysis to laboratory-confirmed infections and SARs from unvaccinated index cases to unvaccinated household contacts to observe how transmission patterns changed independent of vaccination. Next, we evaluated household SARs by variants that were reported in ≥2 studies regardless of vaccination status and restricted to SARs from unvaccinated index cases to unvaccinated contacts for comparison with SAR estimates from our original analyses of the predominantly wild-type variant [1, 11].

Traditionally, vaccine efficacies for reducing susceptibility and infectiousness are estimated as *VE*_*S,p*_ = 1 − *SAR*_01_/*SAR*_00_ and *VE*_*I,p*_ = 1 − *SAR*_10_/*SAR*_00_ respectively, where *SAR*_*ij*_ is the SAR associated with vaccine status *i* (1=vaccinated, 0=unvaccinated) for the index case and *j* for the household contact [8]. The total vaccine effectiveness is defined as *VE*_*T,p*_ =1 − (1− *VE*_*S,p*_)(1 − *VE*_*I,p*_). We examined SARs by index case vaccination status (unvaccinated, partially vaccinated, fully vaccinated, all) and household contact vaccination status (unvaccinated, partially vaccinated, fully vaccinated, all). The resultant SARs were used to estimate *VE*_*S,p*_, *VE*_*I,p*_, and *VE*_*T,p*_. We created forest plots of SARs by index case vaccination status to all household contacts regardless of vaccination status and restricted to unvaccinated contacts only. We also created forest plots of SARs by contact vaccination status from all index cases regardless of vaccination status and from unvaccinated index cases only. Furthermore, we evaluated SARs by vaccine type and vaccination status for index cases and/or household contacts if reported in ≥2 studies. Finally, we evaluated SARs by variant and vaccination status for index cases and/or household contacts if reported in ≥2 studies.

### Evaluation of Study Quality and Risk of Bias

To assess study quality and risk of bias, we used the same modified version of the Newcastle-Ottawa quality assessment scale used by Fung *et al*. and in our first analysis [11, 12]. Studies received up to 9 points based on participant selection (4 points), study comparability (1 point), and outcome of interest (4 points). Studies were classified as having high (≤3 points), moderate (4-6 points), and low (≥7 points) risk of bias. When at least 10 studies were available, we also used funnel plots and Begg and Mazumdar’s rank correlation to evaluate publication bias with significance set at *P*<0.10 [13].

### Statistical Analysis

We used generalized linear mixed-effects models to obtain SAR estimates and 95% CIs. For comparisons across covariate subgroups (variant, index case vaccination status, household contact vaccination status, vaccine type), study was treated as a random effect and the covariate as a fixed effect moderator. For analyses of SARs by index case vaccination status and contact vaccination status, comparisons between subgroups (e.g., fully vaccinated versus unvaccinated index cases) were restricted to pairwise analyses (studies in which SARs were reported from both fully vaccinated and unvaccinated index cases). For vaccine effectiveness measures, we also used generalized linear mixed models to obtain the logit of the SAR and corresponding sampling variances, which were back-transformed to obtain VE summary estimates and 95%CIs. Heterogeneity was measured using the *I*^*2*^ statistic, with thresholds of 25%, 50%, and 75% indicating low, moderate, and high heterogeneity, respectively. All analyses were performed using the metafor package in R software, version 4.1.2 (R Foundation for Statistical Computing) [14, 15]. Statistical significance was set at a 2-tailed *P-*value≤0.05.

## Results

We identified 1,291 records (1,281 from PubMed and 10 from reference lists of eligible articles) published between June 18, 2021 and January 7, 2022 (S1 Figure). Forty-eight new studies [16-64] were included in this review (S2 Table), 4 of which were preprints in our previous review that were subsequently published [53-55, 64].

Forty-nine new studies[16-64] were combined with 77 studies from our previous review [1] for our analysis of household SAR by study period (5 studies were excluded that did not include laboratory-confirmed infections and 1 that included only asymptomatic index cases), resulting in 126 total studies representing 1,437,696 contacts from 35 countries (see S3 Table for references). Figure 1 demonstrates large heterogeneity in SAR over time but estimates with midpoints after July 2020 are generally higher than the earliest estimates. The household SAR for 27 studies with midpoints in 2021 was 35.8% (95%CI, 30.6%-41.3%), whereas the household SAR for 62 studies with midpoints through April 2020 was 15.7% (95%CI, 13.3%-18.4%). Begg and Mazumdar’s rank correlation was statistically significant for studies in 2021 (*P*=0.001), but not studies through April 2020 (S2 Figure). Excluding one study in 2021 [54] that had a relatively low SAR improved the funnel plot symmetry and resulted in a SAR of 33.9% (95%CI, 29.4%-38.7%) for studies with midpoints in 2021. When restricting to unvaccinated contacts only, the household SAR for studies with midpoints in 2021 was 35.4% (95%CI, 30.0%-41.2%).

**Figure 1.**
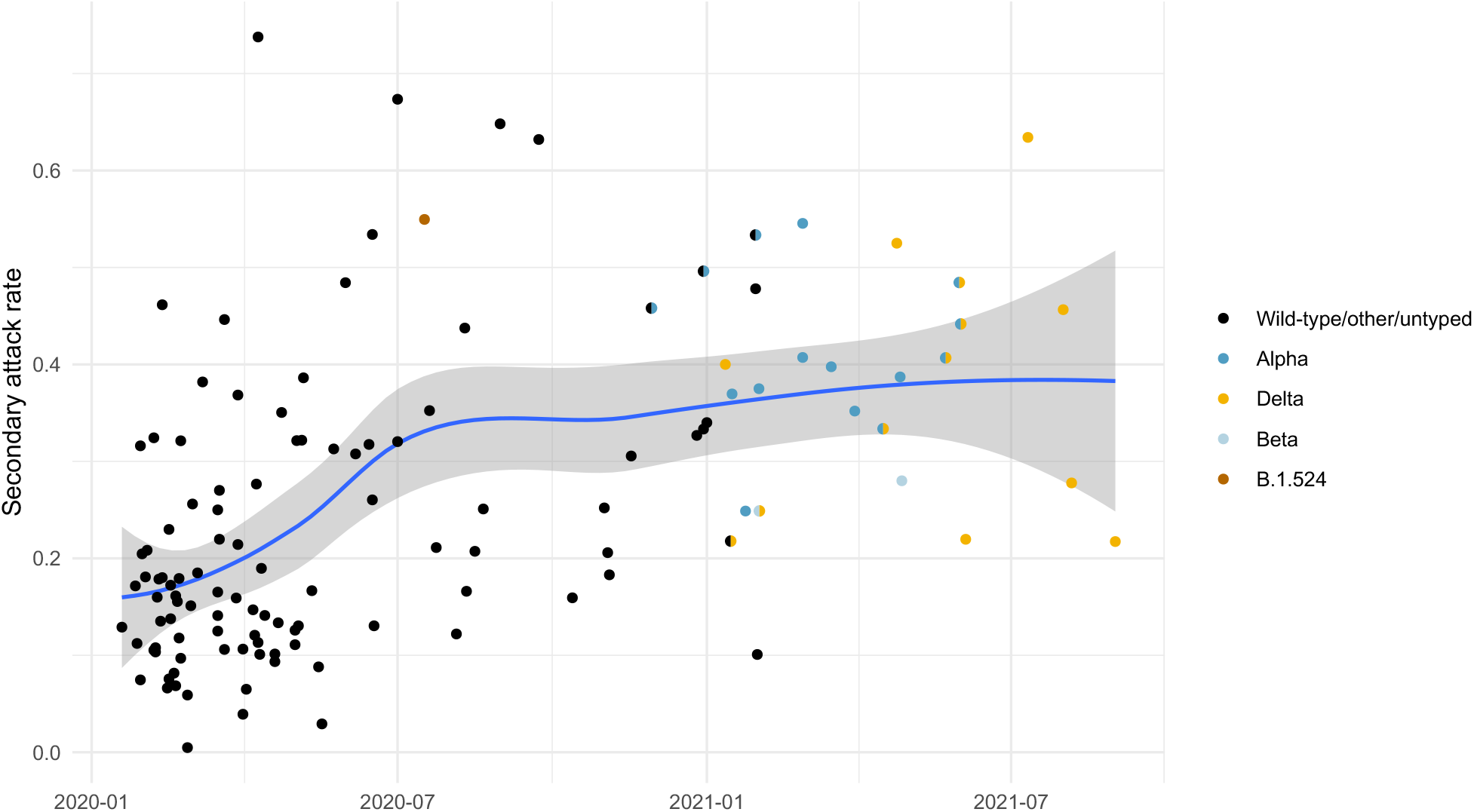
Household secondary attack rates over time (by study midpoint), 126 studies (unvaccinated index cases, unvaccinated contacts). Restricted to laboratory-confirmed results only. The blue line is a loess smoothing line and bands are 95% confidence intervals. Bicolored points represent studies with 2 predominant variants.

Eight new studies [23, 28, 31, 39, 50, 52, 55, 56] were combined with 1 study [65] from our previous review for our analysis of Alpha variant. Figure 2 summarizes results from these 9 studies as well as 12 [23, 25, 29, 32, 41, 43, 48, 52, 56, 61-63] and 3 [20, 23, 56] new studies reporting household SARs for Delta and Beta variants, respectively. Estimated mean household SAR for Alpha was 38.0% (95%CI, 36.0%-40.0%), Delta was 30.8% (95%CI, 23.5%-39.3%), and Beta was 22.5% (95%CI, 18.6%-26.8%) (Figure 2). SARs between Alpha/Delta and Delta/Beta were not significantly different, but Alpha was significantly higher than Beta (*P*<0.001). High heterogeneity was found among studies for Delta (98.0%), and low for Alpha (16.7%) and Beta (2.6%). Begg correlation was not significant for studies of Delta (S3 Figure). SARs did not significantly change for Alpha (37.8%, 95%CI, 35.7%-39.9%) (5 studies) [23, 28, 31, 50, 55] or Delta (27.0%, 95%CI, 18.5%-37.4%) (7 studies) [23, 29, 32, 43, 48, 61, 62] when restricting to studies with low risk of bias (S4 Table). Restricting to unvaccinated contacts only, mean estimated SAR for Delta was 34.9% (95%CI, 26.7%-44.1%) (S4 Figure).

**Figure 2.**
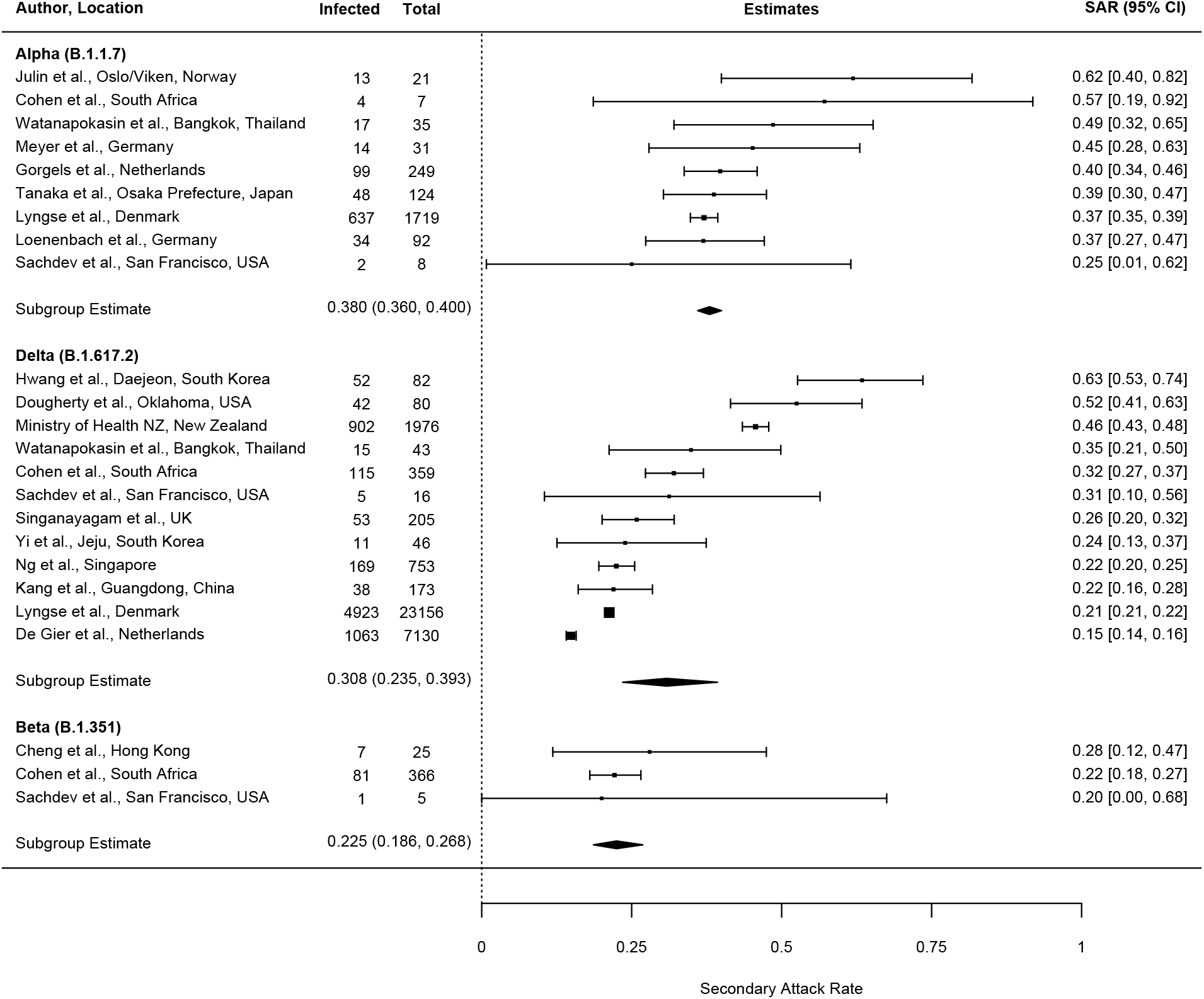
Household secondary attack rates for Alpha (B.1.1.7), Delta (B.1.617.2), and Beta (B.1.351) variants.

Eight studies [24, 26, 34, 39, 43, 48, 54, 62] reported SARs by vaccination status of the index case to all household contacts regardless of vaccination status, seven of which were at low risk of bias and one was moderate (Figure 3). Overall estimated mean SAR was 26.6% (95%CI, 18.7%-36.4%) from unvaccinated (8 studies) [24, 26, 34, 39, 43, 48, 54, 62], 16.2% (95%CI, 8.3%-29.4%) from partially vaccinated (5 studies) [24, 43, 48, 54, 62], and 14.4% (95% CI: 10.5%-19.4%) from fully vaccinated (7 studies) [24, 26, 34, 39, 43, 48, 62] index cases to household contacts. For 7 paired studies [24, 26, 34, 39, 43, 48, 62], estimated mean SAR from unvaccinated index cases (29.9%; 95%CI, 23.0%-37.7%) was significantly higher than from fully vaccinated index cases (*P*<0.001). For 5 paired studies [24, 43, 48, 54, 62], SARs were not significantly different from unvaccinated index cases (19.7%; 95%CI, 13.9%-27.3%) than from partially vaccinated index cases. Three studies included only Delta infections [43, 48, 62]. Restricting to those 3 studies, we found no significant difference in SAR by index case vaccination status to all contacts regardless of vaccination status. Excluding those 3 studies, the estimated mean SAR was significantly higher from unvaccinated index cases (36.3%; 95%CI, 31.3%-41.6%) than from fully vaccinated index cases (10.7%; 95%CI, 9.0%-12.8%) (*P*<0.001) (4 paired studies) [24, 26, 34, 39], but not from partially vaccinated index cases (2 paired studies) [24, 54]. Restricting to unvaccinated household contacts, SARs were also significantly higher from unvaccinated index cases (30.9%, 95%CI, 23.9%-38.8%) than from fully vaccinated index cases (12.0%, 95%CI, 10.0%-14.2%) (*P*<0.001) (4 paired studies) [24, 26, 48, 62], but not from partially vaccinated index cases (3 paired studies) [24, 54, 62] (S5 Figure). SARs were generally lower from fully vaccinated index cases regardless of contact vaccination status (S6 Figure). Direct comparison of these studies is compromised, however, because of differences between studies in terms of vaccine types, definition of vaccination status (e.g., time elapsed since vaccination or dosage) (S5 Table), vaccination coverage among contacts, characteristics of the study population, duration of follow-up, diagnostic procedures and tools, location, magnitude of the pandemic, and circulating variants.

**Figure 3.**
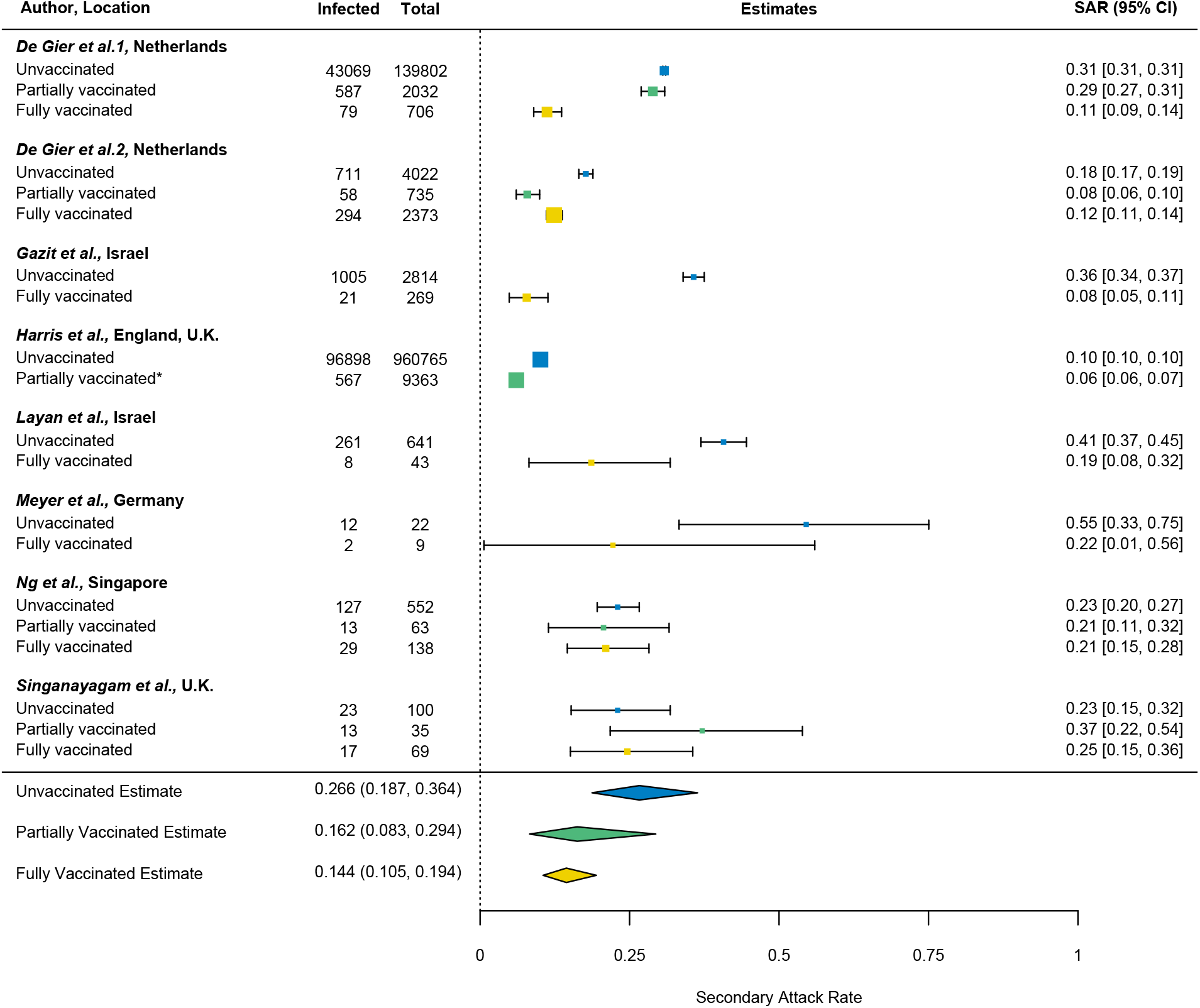
Household secondary attack rates by index case vaccination status. All contacts are included regardless of vaccination status. *For Harris *et al*., most of the vaccinated index cases (93%) had received only the first dose of vaccine and secondary attack rates were not disaggregated by dose.

Figure 4 summarizes 9 studies [24, 26, 34, 38, 43, 48, 56, 62, 63] reporting household SARs by contact vaccination status regardless of index case vaccination status, eight of which were at low risk of bias and two were moderate. Overall estimated mean SAR was 33.8% (95%CI, 28.0%-40.2%) to unvaccinated contacts (9 studies) [24, 26, 34, 38, 43, 48, 56, 62, 63], 23.7% (95%CI, 19.1%-28.9%) to partially vaccinated contacts (6 studies) [24, 38, 43, 48, 56, 63], and 14.1% (95%CI, 10.6%-18.6%) to fully vaccinated contacts (9 studies) [24, 26, 34, 38, 43, 48, 56, 62, 63]. In the 9 paired studies, estimated mean household SARs were significantly higher to unvaccinated contacts than to fully vaccinated contacts (*P*<0.001). For 6 paired studies [24, 38, 43, 48, 56, 63], SARs were significantly higher to unvaccinated contacts (33.1%, 95%CI, 27.8%-38.8%) than to partially vaccinated contacts (*P=*0.020), but SARs were not significantly different to partially vaccinated contacts than to fully vaccinated contacts (16.6%, 95%CI, 11.9%-22.9%). SARs were consistent when restricting to only unvaccinated index cases (4 studies [24, 26, 48, 62]) (S7 Figure). When restricting to 4 studies [43, 48, 62, 63] that targeted Delta, SARs were also significantly higher to unvaccinated contacts (24.4%, 95%CI, 19.3%-30.4%) than to fully vaccinated contacts (14.3%, 95%CI, 9.3%-21.3%) (*P*=0.027). We also estimated vaccine effectiveness based on the SARs (Table 1).

**Table 1.** Estimated vaccine effectiveness (95%CI) estimates from household secondary attack rates.

|  |  |  |
| --- | --- | --- |
| Vaccine effectiveness | Full Vaccination | Partial Vaccination |
| $VE_{I,p}$ | 56.6% (28.7%-73.6%) | 27.5% (-6.4%-50.7%) |
| $VE_{S,p}$ | 70.3% (59.3%-78.4%) | 43.9% (21.8%-59.7%) |
| $VE_{T,p}$ | 86.8% (76.7%-92.5%) | 59.9% (34.4%-75.5%) |
| $VE_{I,p}$: vaccine effectiveness for infectiousness based on the transmission probability $p$; $VE_{S,p}$: vaccine effectiveness for susceptibility; $VE_{T,p}$: total vaccine effectiveness | | |

**Figure 4.**
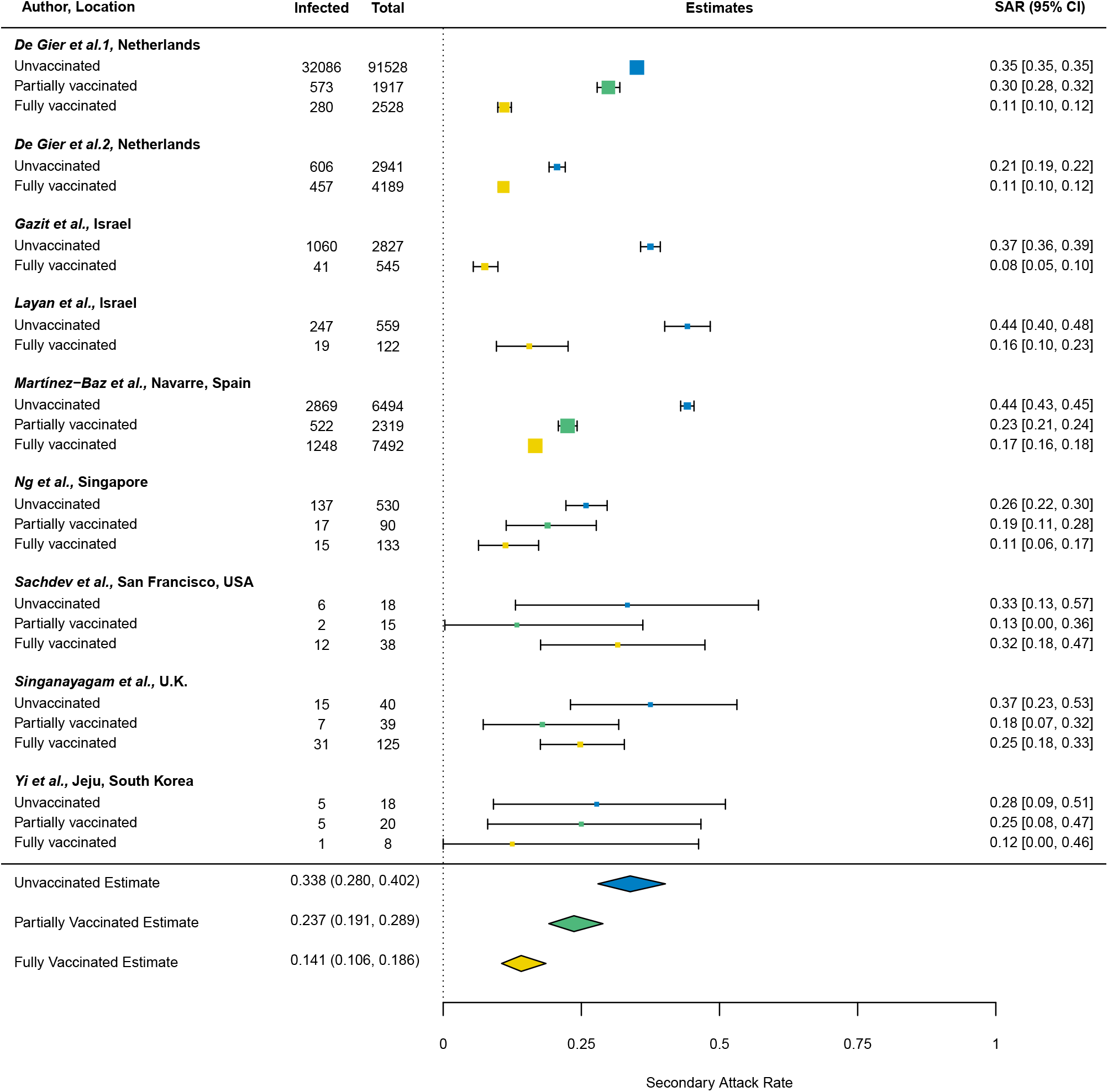
Household secondary attack rates by contact vaccination status. All index cases are included regardless of vaccination status.

Next, we examined SARs by vaccine type and index case vaccination status regardless of vaccination status of household contacts. SARs were included in 2 studies [26, 39] for BNT162b2 and the mean estimated SAR from fully vaccinated index cases was 8.3% (95%CI, 5.6%-12.1%) compared to 35.9% from unvaccinated index cases (95%CI, 34.1%-37.6%) (S8 Figure).

We also examined SARs by vaccine type and contact vaccination status regardless of index case vaccination status (3 studies [24, 38, 56]). Mean estimated SAR to household contacts fully vaccinated with Ad26.COV2.S (1 dose) (42.7%, 95% CI: 13.6%-77.9%) (*P*=0.005) or BNT162b2 (15.8%, 95%CI, 15.0%-16.7%) (*P*<0.001) was significantly higher than to contacts fully vaccinated with mRNA-1273 (2 doses) (6.2%, 95% CI: 2.8%-13.0%) (S9 Figure). Additionally, mean estimated SAR was higher to contacts partially vaccinated with ChAdOx1-S (29.5%, 95% CI: 24.0%-35.7%) than contacts partially vaccinated with mRNA-1273 (17.5%, 95%CI, 13.7%-22.3%) (*P*=0.008). There was no significant difference in SAR to contacts fully vaccinated for ChAdOx1-S and BNT162b2, Ad26.COV2.S, or mRNA-1273; or to contacts partially vaccinated for BNT162b2 and mRNA-1273or ChAdOx1-S.

## Discussion

We aggregated household studies to examine how variants of concern and vaccination affected household transmission rates of SARS-CoV-2. Household SARs from fully vaccinated index cases were lower than from unvaccinated index cases. Fully and partially vaccinated household contacts were less susceptible to SARS-CoV-2 infection than unvaccinated contacts. SARs for Delta and Alpha were significantly higher than estimates for the original wild-type variant.

Several individual studies included in this analysis reported that full vaccination of index cases significantly reduced the risk of transmission to household contacts [24, 34]. Conversely, other studies included in this analysis reported that vaccination status of the index case was not associated with household contact infection [43, 48]. A meta-analysis allows us to aggregate all the evidence of index case vaccination status from multiple studies and control for differences between the studies. We found lower transmission to household contacts from fully vaccinated index cases than from unvaccinated index cases, but not from partially vaccinated index cases. An observational cohort study from England which included contacts outside the household also reported that two doses of BNT162b2 or ChAdOx1 reduced onward transmission of Delta, but by less than Alpha, and the impact of vaccination against onward transmission waned over time [66]. Our estimate for *VE*_*I,p*_ of 56.6% was within the 41%-79% range reported for *VE*_*I*_ from a modeling study that used household data from Israel before Delta became widespread [67]. Potential mechanisms for reduced infectiousness following vaccination include decreases in the respiratory tract viral load and severity of symptoms [68].

Fully vaccinated and partially vaccinated contacts had significantly lower SARs than unvaccinated contacts. Other observational studies demonstrated reduced susceptibility to infection among high risk or household contacts vaccinated with BNT162b2 or ChAdOx1 in Scotland [69], BNT162b2 in Sweden [70], and BNT162b2 or mRNA-1273 in Belgium [71]. Studies have reported that full vaccination with mRNA vaccines or ChAdOx1 effectively prevent infection against the original wild-type, Alpha, and Beta variants, but are less protective against infection for Delta [72, 73]. Our estimates of *VE*_*S,p*_ (70.3%, 95%CI, 59.3%-78.4%) and *VE*_*T,p*_ (86.8%, 95%CI, 76.7%-92.5%) were slightly lower than the age-adjusted *VE*_*S*_ (80.5%, 95%CI, 78.9%-82.1%) and *VE*_*T*_ (88.5%, 95%CI, 82.3%-94.8%) reported by *Prunas et al* [67]. Myriad factors preclude our ability to make direct comparisons of vaccine effectiveness across studies including differences in the study population (e.g., age, comorbidities, serostatus), viral characteristics, vaccine type, time period defining vaccination status, intensity of the epidemic, community behavior, and use of nonpharmaceutical interventions (masks, social distancing) [74]. For example, in this analysis *Singanayagam et al*. [48] included households of any size with contacts ≥5 years, whereas *Gazit et al*. [26] restricted to households with only one contact other than the index case. Moreover, *Ng et al*. in Singapore reported that all identified close contacts were placed under a legally-binding quarantine for 14 days during which they were not allowed to leave their homes [43], whereas contacts in other studies may have had a higher risk of infection outside the household.

With the addition of 49 studies since our last review [1], we observed higher SARs in 2021 than earlier in the pandemic. This pattern may be attributed in part to the emergence of more contagious variants. SAR estimates for Alpha (38.0%) and Delta (30.8%) variants were both higher than the overall SAR previously reported (18.9%) for study periods earlier in the pandemic when the wild-type variant was prevalent [1]. Public Health England (PHE), which tracks SARs for variants of concern and variants of interest regardless of vaccination status for index cases and household contacts, found SARs similar for Alpha (10.2%, 95%CI, 10.1%-10.3%) and Delta (10.4%, 95%CI, 10.4%-10.5%) variants [75]. They note, however, that direct comparisons between variants are not valid as vaccination levels and social restrictions in England have varied over this period. Similarly, SARs for Delta and Alpha were not significantly different in this study even when restricting to unvaccinated contacts only, which may be partially attributed to an increase in population immunity consequent to infection. A prospective cohort study [22] and case-control study [76] in England demonstrated increased household transmission for Delta compared to Alpha. Increased transmissibility may be attributed to higher viral loads, shorter incubation periods, and mutations in the spike glycoprotein of the virus, which may confer immune escape potential [77]. Delta infections produced more viral RNA copies per mL than Alpha infections [78], its *in vitro* replication rate is higher than Alpha [79], and its spike protein binds more efficiently to the host cell entry receptor ACE2 protein [80].

There was large heterogeneity in SARs over time which may be attributed to variations in study methods, environmental factors, and contact patterns. Comparisons of SARs by vaccination status between studies were also hindered by differences between studies and there were few studies disaggregating SARs by both vaccination status of the index cases and contacts. The studies included in this review are from contact tracing investigations which are more likely to identify symptomatic index cases than asymptomatic individuals, which could inflate the crude SAR. This may also underestimate the reduction in transmission from vaccination for people infected with Delta [81]. Our analyses by vaccine type and Beta variant were limited to three studies. There was insufficient data to determine vaccine effectiveness for specific subgroups (e.g., by age group) and whether that varied by variant.

Household contacts exposed to Delta or Alpha variants are at increased risk of infection compared to the original wild-type variant from Wuhan. Vaccination was demonstrated to reduce susceptibility to infection and infectiousness. The household remains an important venue of transmission for SARS-CoV-2. Other public health measures such as hygiene, increased testing, isolation, and improved ventilation may help limit its spread. Preliminary analyses from PHE demonstrate increased odds of household transmission from Omicron index cases than from Delta index cases, adjusting for index case vaccination status and other factors [82]. A study from Denmark reported higher transmission rates for Omicron than Delta for fully vaccinated individuals but not unvaccinated individuals [61]. The transmissibility and virulence of Omicron is only now being elucidated and other variants are likely to emerge.

## Data Availability

All relevant data are within the manuscript.

## Acknowledgments

This work was supported by the National Institutes of Health [R01-AI139761 to ZJM, YY, IML, MEH, NED].

## References

1. Madewell ZJ, Yang Y, Longini IM, Jr., Halloran ME, Dean NE. Factors Associated With Household Transmission of SARS-CoV-2: An Updated Systematic Review and Meta-analysis. JAMA Netw Open 2021; 4(8): e2122240.

2. Centers for Disease Control and Prevention. Global Variants Report. Available at: https://covid.cdc.gov/covid-data-tracker/#global-variant-report-map.

3. Willett BJ, Grove J, MacLean O, et al. The hyper-transmissible SARS-CoV-2 Omicron variant exhibits significant antigenic change, vaccine escape and a switch in cell entry mechanism. medRxiv 2022.

4. Shapiro J, Dean NE, Madewell ZJ, Yang Y, Halloran ME, Longini I. Efficacy Estimates for Various COVID-19 Vaccines: What we Know from the Literature and Reports. medRxiv 2021: 2021.05.20.21257461.

5. Moustsen-Helms IR, Emborg H-D, Nielsen J, et al. Vaccine effectiveness after 1st and 2nd dose of the BNT162b2 mRNA Covid-19 Vaccine in long-term care facility residents and healthcare workers – a Danish cohort study. medRxiv 2021: 2021.03.08.21252200.

6. Haas EJ, Angulo FJ, McLaughlin JM, et al. Impact and effectiveness of mRNA BNT162b2 vaccine against SARS-CoV-2 infections and COVID-19 cases, hospitalisations, and deaths following a nationwide vaccination campaign in Israel: an observational study using national surveillance data. Lancet 2021; 397(10287): 1819–29.

7. Lopez Bernal J, Andrews N, Gower C, et al. Effectiveness of Covid-19 vaccines against the B. 1.617. 2 (Delta) variant. N Engl J Med 2021: 585–94.

8. Halloran ME, Longini IM, Struchiner CJ, Longini IM. Design and analysis of vaccine studies: Springer, 2010.

9. Datta S, Halloran ME, Longini IM, Jr. Efficiency of estimating vaccine efficacy for susceptibility and infectiousness: randomization by individual versus household. Biometrics 1999; 55(3): 792–8.

10. Préziosi MP, Halloran ME. Effects of pertussis vaccination on transmission: vaccine efficacy for infectiousness. Vaccine 2003; 21(17-18): 1853–61.

11. Madewell ZJ, Yang Y, Longini IM, Jr., Halloran ME, Dean NE. Household Transmission of SARS-CoV-2: A Systematic Review and Meta-analysis. JAMA Netw Open 2020; 3(12): e2031756.

12. Fung HF, Martinez L, Alarid-Escudero F, et al. The household secondary attack rate of severe acute respiratory syndrome coronavirus 2 (SARS-CoV-2): a rapid review. Clinical Infectious Diseases 2021; 73(Supplement_2): S138–S45.

13. Begg CB, Mazumdar M. Operating characteristics of a rank correlation test for publication bias. Biometrics 1994: 1088–101.

14. Viechtbauer W. Conducting meta-analyses in R with the metafor package. Journal of statistical software 2010; 36(3): 1–48.

15. R Core Team. R: A language and environment for statistical computing. Available at: http://r.meteo.uni.wroc.pl/web/packages/dplR/vignettes/intro-dplR.pdf.

16. Afonso ET, Marques SM, Costa LD, et al. Secondary household transmission of SARSDCoVD2 among children and adolescents: Clinical and epidemiological aspects. Pediatric Pulmonology 2021.

17. Bistaraki A, Roussos S, Tsiodras S, Sypsa V. Age-dependent effects on infectivity and susceptibility to SARS-CoV-2 infection: results from nationwide contact tracing data in Greece. Infectious Diseases 2021: 1–10.

18. Burke RM, Calderwood L, Killerby ME, et al. Patterns of Virus Exposure and Presumed Household Transmission among Persons with Coronavirus Disease, United States, January–April 2020. Emerging Infectious Diseases 2021; 27(9): 2323.

19. Calvani M, Cantiello G, Cavani M, et al. Reasons for SARS-CoV-2 infection in children and their role in the transmission of infection according to age: a case-control study. Ital J Pediatr 2021; 47(1): 193.

20. Cheng VC-C, Siu GK-H, Wong S-C, et al. Complementation of contact tracing by mass testing for successful containment of beta COVID-19 variant (SARS-CoV-2 VOC B. 1.351) epidemic in Hong Kong. The Lancet Regional Health-Western Pacific 2021; 17: 100281.

21. Chu VT, Yousaf AR, Chang K, et al. Household transmission of SARS-CoV-2 from children and adolescents. New England Journal of Medicine 2021; 385(10): 954–6.

22. Clifford S, Waight P, Hackman J, et al. Effectiveness of BNT162b2 and ChAdOx1 against SARS-CoV-2 household transmission: a prospective cohort study in England. medRxiv 2021: 2021.11.24.21266401.

23. Cohen C, Kleynhans J, von Gottberg A, et al. SARS-CoV-2 incidence, transmission and reinfection in a rural and an urban setting: results of the PHIRST-C cohort study, South Africa, 2020-2021. medRxiv 2021: 2021.07.20.21260855.

24. de Gier B, Andeweg S, Joosten R, et al. Vaccine effectiveness against SARS-CoV-2 transmission and infections among household and other close contacts of confirmed cases, the Netherlands, February to May 2021. Eurosurveillance 2021; 26(31): 2100640.

25. Dougherty K, Mannell M, Naqvi O, Matson D, Stone J. SARS-CoV-2 B. 1.617. 2 (Delta) variant COVID-19 outbreak associated with a gymnastics facility—Oklahoma, April–May 2021. Morbidity and Mortality Weekly Report 2021; 70(28): 1004.

26. Gazit S, Mizrahi B, Kalkstein N, et al. BNT162b2 mRNA Vaccine Effectiveness Given Confirmed Exposure: Analysis of Household Members of COVID-19 Patients. Clinical Infectious Diseases 2021.

27. Ge Y, Martinez L, Sun S, et al. COVID-19 transmission dynamics among close contacts of index patients with COVID-19: a population-based cohort study in Zhejiang province, China. JAMA Internal Medicine 2021; 181(10): 1343–50.

28. Gorgels K, Alphen L, Hackert BvdVV, et al. Increased Transmissibility of SARS-CoV-2 Alpha Variant (B.1.1.7) in Children: Three Large Primary School Outbreaks Revealed by Whole Genome Sequencing in the Netherlands. Research Square 2021.

29. Hwang H, Lim J-S, Song S-A, et al. Transmission dynamics of the Delta variant of SARS-CoV-2 infections in South Korea. The Journal of Infectious Diseases 2021.

30. Jagdale GR, Parande MA, Borle P, et al. Secondary Attack Rate among the Contacts of COVID-19 Patients at the Beginning of the Pandemic in Pune City of Western Maharashtra, India. Journal of Communicable Diseases (E-ISSN: 2581-351X & P-ISSN: 0019-5138) 2021; 53(3): 51–9.

31. Julin CH, Robertson AH, Hungnes O, et al. Household Transmission of SARS-CoV-2: A Prospective Longitudinal Study Showing Higher Viral Load and Increased Transmissibility of the Alpha Variant Compared to Previous Strains. Microorganisms 2021; 9(11): 2371.

32. Kang M, Xin H, Yuan J, et al. Transmission dynamics and epidemiological characteristics of Delta variant infections in China. Medrxiv 2021.

33. Karumanagoundar K, Raju M, Ponnaiah M, et al. Secondary attack rate of COVID-19 among contacts and risk factors, Tamil Nadu, March–May 2020: a retrospective cohort study. BMJ open 2021; 11(11): e051491.

34. Layan M, Gilboa M, Gonen T, et al. Impact of BNT162b2 vaccination and isolation on SARS-CoV-2 transmission in Israeli households: an observational study. MedRxiv 2021.

35. Li Y, Liu J, Yang Z, et al. Transmission of Severe Acute Respiratory Syndrome Coronavirus 2 to Close Contacts, China, January–February 2020. Emerging Infectious Diseases 2021; 27(9): 2288.

36. Liu PY, Gragnani CM, Timmerman J, et al. Pediatric Household Transmission of Severe Acute Respiratory Coronavirus-2 Infection—Los Angeles County, December 2020 to February 2021. The Pediatric Infectious Disease Journal 2021; 40(10): e379.

37. Martinez DA, Klein EY, Parent C, et al. Latino Household Transmission of Severe Acute Respiratory Syndrome Coronavirus 2. Clinical Infectious Diseases 2021.

38. Martínez-Baz I, Trobajo-Sanmartín C, Miqueleiz A, et al. Product-specific COVID-19 vaccine effectiveness against secondary infection in close contacts, Navarre, Spain, April to August 2021. Eurosurveillance 2021; 26(39): 2100894.

39. Meyer ED, Sandfort M, Bender J, et al. Two doses of the mRNA BNT162b2 vaccine reduce severe outcomes, viral load and secondary attack rate: evidence from a SARS-CoV-2 Alpha outbreak in a nursing home in Germany, January-March 2021. medRxiv 2021: 2021.09.13.21262519.

40. Miller E, Waight PA, Andrews NJ, et al. Transmission of SARS-CoV-2 in the household setting: A prospective cohort study in children and adults in England. Journal of Infection 2021; 83(4): 483–9.

41. Ministry of Health NZ. COVID-19 Variants Update. Available at: https://www.health.govt.nz/system/files/documents/pages/22-november-2021-variants-update-summary.pdf.

42. Musa S, Kissling E, Valenciano M, et al. Household transmission of SARS-CoV-2: a prospective observational study in Bosnia and Herzegovina, August–December 2020. International Journal of Infectious Diseases 2021; 112: 352–61.

43. Ng OT, Koh V, Chiew CJ, et al. Impact of Delta Variant and Vaccination on SARS-CoV-2 Secondary Attack Rate Among Household Close Contacts. The Lancet Regional Health-Western Pacific 2021; 17: 100299.

44. Ng DCE, Tan KK, Chin L, et al. Risk factors associated with household transmission of SARSDCoVD2 in Negeri Sembilan, Malaysia. Journal of paediatrics and child health 2021.

45. Ogata T, Irie F, Ogawa E, et al. Secondary Attack Rate among Non-Spousal Household Contacts of Coronavirus Disease 2019 in Tsuchiura, Japan, August 2020–February 2021. International Journal of Environmental Research and Public Health 2021; 18(17): 8921.

46. Rajmohan P, Jose P, Thodi JBA, et al. Dynamics of transmission of COVID-19 cases and household contacts: A prospective cohort study. Journal of Acute Disease 2021; 10(4): 162.

47. Ratovoson R, Razafimahatratra R, Randriamanantsoa L, et al. Household transmission of COVID-19 among the earliest cases in Antananarivo, Madagascar. Influenza Other Respir Viruses 2021.

48. Singanayagam A, Hakki S, Dunning J, et al. Community transmission and viral load kinetics of the SARS-CoV-2 delta (B. 1.617. 2) variant in vaccinated and unvaccinated individuals in the UK: a prospective, longitudinal, cohort study. The Lancet Infectious Diseases 2021.

49. Soriano-Arandes A, Gatell A, Serrano P, et al. Household Severe Acute Respiratory Syndrome Coronavirus 2 Transmission and Children: A Network Prospective Study. Clin Infect Dis 2021; 73(6): e1261–e9.

50. Tanaka H, Hirayama A, Nagai H, et al. Increased transmissibility of the SARS-CoV-2 alpha variant in a japanese population. International Journal of Environmental Research and Public Health 2021; 18(15): 7752.

51. ur Rehman S, Qaisrani M, Abbasi S, et al. COVID-19 outbreak in Islamabad resulting from a travel-associated primary case: A case series. Global Biosecurity 2021; 3(1).

52. Watanapokasin N, Siripongboonsitti T, Ungtrakul T, et al. Transmissibility of SARS-CoV-2 variants as a secondary attack in Thai households: a retrospective study. IJID Regions 2021; 1: 1–2.

53. Cerami C, Popkin-Hall ZR, Rapp T, et al. Household transmission of SARS-CoV-2 in the United States: living density, viral load, and disproportionate impact on communities of color. Clin Infect Dis 2021.

54. Harris RJ, Hall JA, Zaidi A, Andrews NJ, Dunbar JK, Dabrera G. Effect of Vaccination on Household Transmission of SARS-CoV-2 in England. New England Journal of Medicine 2021.

55. Lyngse FP, Mølbak K, Skov RL, et al. Increased transmissibility of SARS-CoV-2 lineage B.1.1.7 by age and viral load. Nature Communications 2021; 12(1): 7251.

56. Sachdev DD, Chew Ng R, Sankaran M, et al. Contact tracing outcomes among household contacts of fully vaccinated COVID-19 patients -San Francisco, California, January 29-July 2, 2021. Clin Infect Dis 2021.

57. Dub T, Nohynek H, Hagberg L, et al. High secondary attack rate and persistence of SARS-CoV-2 antibodies in household transmission study participants, Finland 2020. 2021.

58. Montecucco A, Dini G, Rahmani A, et al. Investigating SARS-CoV-2 transmission among co-workers in a University of Northern Italy during COVID-19 pandemic: an observational study. La Medicina del Lavoro | Work, Environment and Health 2021; 112(6): 429–35.

59. Remón-Berrade M, Guillen-Aguinaga S, Sarrate-Adot I, et al. Risk of Secondary Household Transmission of COVID-19 from Health Care Workers in a Hospital in Spain. Epidemiologia 2022; 3(1): 1–10.

60. Loss J, Wurm J, Varnaccia G, et al. Transmission of SARS-CoV-2 among children and staff in German daycare centers: results from the COALA study. medRxiv 2021: 2021.12.21.21268157.

61. Lyngse FP, Mortensen LH, Denwood MJ, et al. SARS-CoV-2 Omicron VOC Transmission in Danish Households. medRxiv 2021: 2021.12.27.21268278.

62. de Gier B, Andeweg S, Backer JA, et al. Vaccine effectiveness against SARS-CoV-2 transmission to household contacts during dominance of Delta variant (B.1.617.2), August-September 2021, the Netherlands. medRxiv 2021: 2021.10.14.21264959.

63. Yi S, Kim JM, Choe YJ, et al. SARS-CoV-2 Delta Variant Breakthrough Infection and Onward Secondary Transmission in Household. J Korean Med Sci 2022; 37(1): 0.

64. Tanaka ML, Marentes Ruiz CJ, Malhotra S, et al. SARS-CoV-2 Transmission Dynamics in Households With Children, Los Angeles, California. Frontiers in Pediatrics 2022; 9(1520).

65. Loenenbach A, Markus I, Lehfeld A-S, et al. SARS-CoV-2 variant B. 1.1. 7 susceptibility and infectiousness of children and adults deduced from investigations of childcare centre outbreaks, Germany, 2021. Eurosurveillance 2021; 26(21): 2100433.

66. Eyre DW, Taylor D, Purver M, et al. The impact of SARS-CoV-2 vaccination on Alpha & Delta variant transmission. medRxiv 2021: 2021.09.28.21264260.

67. Prunas O, Warren JL, Crawford FW, et al. Vaccination with BNT162b2 reduces transmission of SARS-CoV-2 to household contacts in Israel. medRxiv 2021: 2021.07.13.21260393.

68. Richterman A, Meyerowitz EA, Cevik M. Indirect Protection by Reducing Transmission: Ending the Pandemic with SARS-CoV-2 Vaccination. Open Forum Infectious Diseases 2021: ofab259.

69. Shah AS, Gribben C, Bishop J, et al. Effect of vaccination on transmission of SARS-CoV-2. New England Journal of Medicine 2021; 385(18): 1718–20.

70. Björk J, Inghammar M, Moghaddassi M, Rasmussen M, Malmqvist U, Kahn F. High level of protection against COVID-19 after two doses of BNT162b2 vaccine in the working age population–first results from a cohort study in Southern Sweden. Infectious Diseases 2021: 1–6.

71. Braeye T, Cornelissen L, Catteau L, et al. Vaccine effectiveness against infection and onwards transmission of COVID-19: Analysis of Belgian contact tracing data, January-June 2021. Vaccine 2021; 39(39): 5456–60.

72. Fiolet T, Kherabi Y, MacDonald C-J, Ghosn J, Peiffer-Smadja N. Comparing COVID-19 vaccines for their characteristics, efficacy and effectiveness against SARS-CoV-2 and variants of concern: A narrative review. Clinical Microbiology and Infection 2021.

73. Seppälä E, Veneti L, Starrfelt J, et al. Vaccine effectiveness against infection with the Delta (B. 1.617. 2) variant, Norway, April to August 2021. Eurosurveillance 2021; 26(35): 2100793.

74. Madewell ZJ, Dean NE, Berlin JA, et al. Challenges of evaluating and modelling vaccination in emerging infectious diseases. Epidemics 2021; 37: 100506.

75. Public Health England. SARS-CoV-2 variants of concern and variants under investigation in England: Technical briefing 23. Available at: https://assets.publishing.service.gov.uk/government/uploads/system/uploads/attachment_data/file/1018547/Technical_Briefing_23_21_09_16.pdf.

76. Allen H, Vusirikala A, Flannagan J, et al. Household transmission of COVID-19 cases associated with SARS-CoV-2 delta variant (B.1.617.2): national case-control study. Lancet Reg Health Eur 2021: 100252.

77. Kumar V, Singh J, Hasnain SE, Sundar D. Possible link between higher transmissibility of alpha, kappa and delta variants of SARS-CoV-2 and increased structural stability of its spike protein and hACE2 affinity. International journal of molecular sciences 2021; 22(17): 9131.

78. Earnest R, Uddin R, Matluk N, et al. Comparative transmissibility of SARS-CoV-2 variants Delta and Alpha in New England, USA. medRxiv 2021: 2021.10.06.21264641.

79. Mlcochova P, Kemp SA, Dhar MS, et al. SARS-CoV-2 B. 1.617. 2 Delta variant replication and immune evasion. Nature 2021: 1–6.

80. Zhang J, Xiao T, Cai Y, et al. Membrane fusion and immune evasion by the spike protein of SARS-CoV-2 Delta variant. Science 2021: eabl9463.

81. Meyerowitz E, Richterman A. SARS-CoV-2 Transmission and Prevention in the Era of the Delta Variant. Available at SSRN 3964247 2021.

82. Public Health England. SARS-CoV-2 variants of concern and variants under investigation in England: Technical briefing 31.

