## Supplement for "Household secondary attack rates of SARS-CoV-2 by variant and vaccination status: an updated systematic review and meta-analysis"

**S1 Figure. PRISMA flow diagram.**

**S2 Figure. Funnel plots of studies reporting household secondary attack rates with midpoints in 2021 and through April 2020.**

**S3 Figure. Household secondary attack rates by index case vaccination status. Only unvaccinated contacts are included.**

**S4 Figure. Household secondary attack rates by vaccination status of the index cases and contacts. The number of studies is provided.**

**S5 Figure. Household secondary attack rates by contact vaccination status. Only unvaccinated index cases are included**

**S6 Figure. Household secondary attack rates for BNT162b2 vaccine by index case vaccination status. All contacts are included regardless of vaccination status.**

**S7 Figure. Household secondary attack rates by vaccine and contact vaccination status. All index cases are included regardless of vaccination status.**

**S1 Table. Electronic databases and search strategy for household secondary attack rate of SARS-CoV-2.**

**S2 Table. Description of studies identified from June 18, 2021 to January 7, 2022.**

**S3 Table. References for studies included in analysis of SAR over time.**

**S4 Table. Risk of bias assessment for studies included in review of household transmissibility of SARS-CoV-2.**

**S5 Table. Vaccination status definitions for studies that reported household secondary attack rates by vaccination status of index cases or contacts.**

**S1 Figure. PRISMA flow diagram**

**Identification of new studies via other methods**

**Identification of new studies via databases and registers**

**Previous studies**

Records removed *before screening*:

Records published through June 17, 2021, the last search date of our previous review (n = 5,578)

Studies included in previous version of review (n = 87 studies)

Records screened on title and abstract (n = 1,287)

Records identified from databases (n = 6,865)

New studies included (n = 49)

- Studies with original data (N=45)
- Preprints in first analysis that were subsequently published (n = 4)

Records identified from reference lists of eligible articles (n = 11)

**Identification**

Records excluded (n = 1,196)

Reports excluded (n = 53):

- No data on uninfected contacts (n = 17)

- No data on household contacts (n = 13)

- Reported prevalence or overall household attack rate, which includes index cases (n = 7)

- Tested household contacts using antibody tests (n = 6)

- Case reports or cluster investigations that focused on individual households or families (n = 3)

- No original data (n = 3)

- No data on transmission (n = 2)

- Overlapping study population with another article included in meta-analysis (n = 2)

**Screening**

Reports assessed for eligibility

(n = 91)

Reviewed methodology to exclude studies that did not include laboratory-confirmed infections and that only included asymptomatic index cases (n = 87)

Records excluded

(n = 6)

**Included**

Total studies included in review

(n = 126)

**S2 Figure. Funnel plots of studies reporting household secondary attack rates with midpoints in 2021 and through April 2020.**

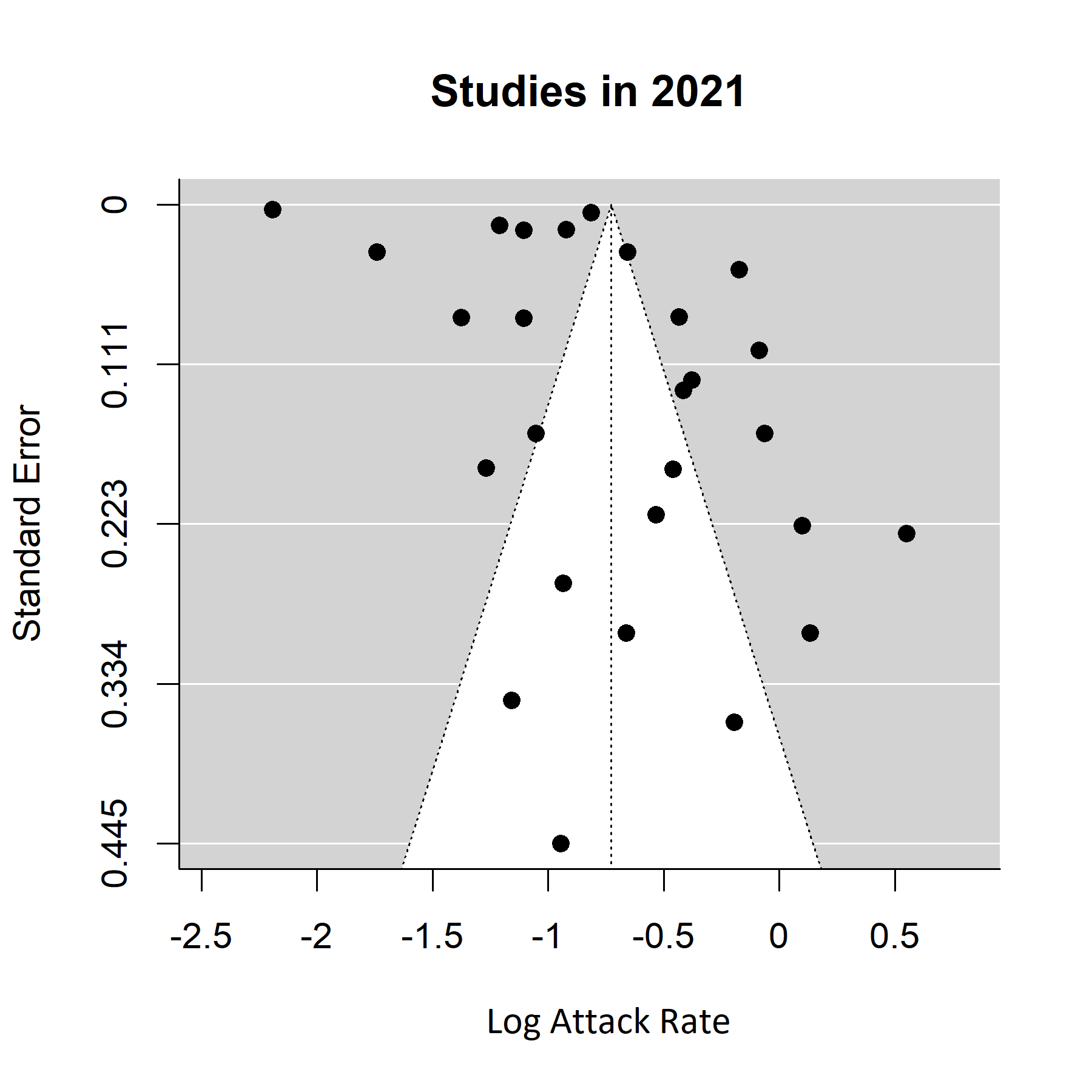

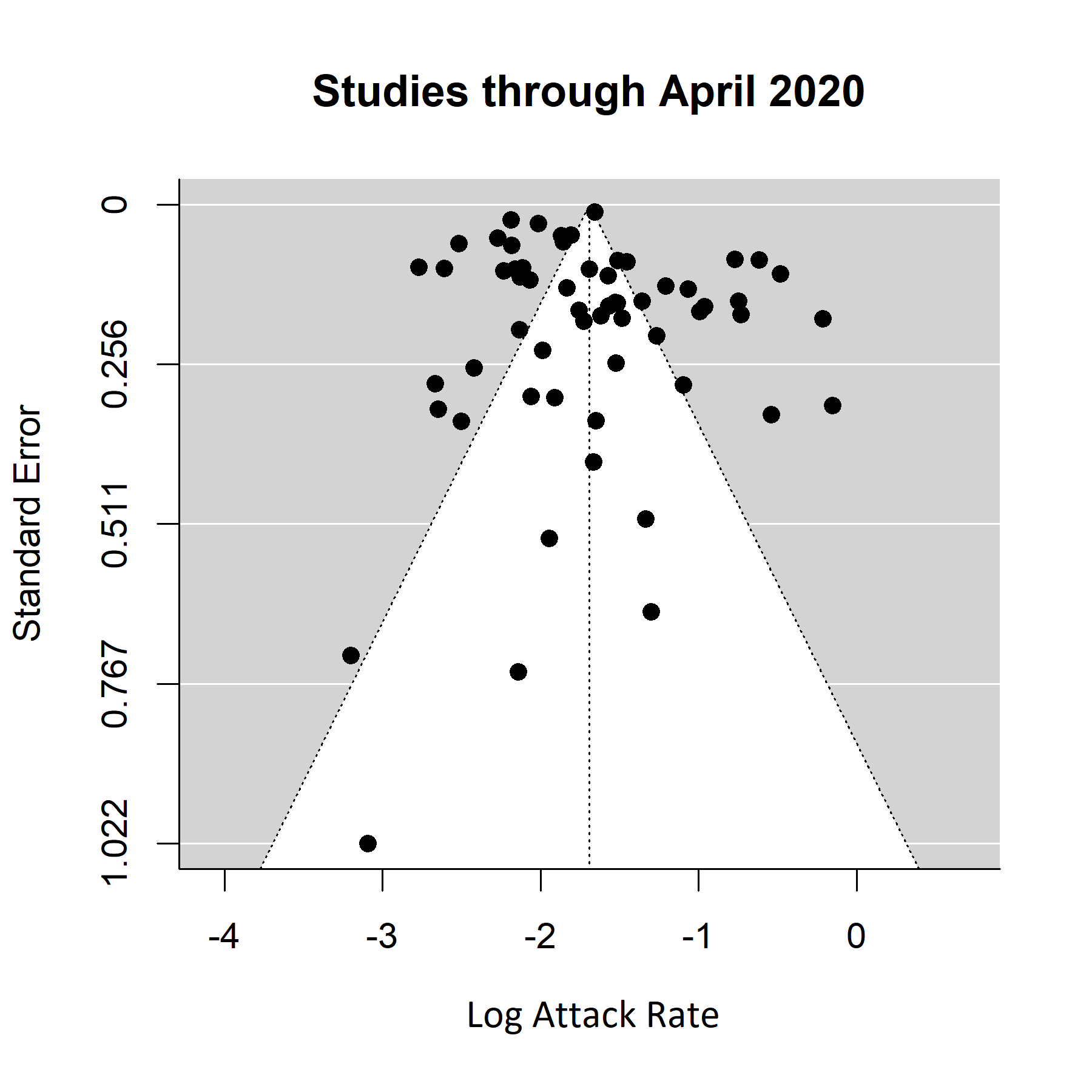

**S3 Figure. Funnel plots of studies reporting household secondary attack rates for Delta (B.1.617.2) variant.**

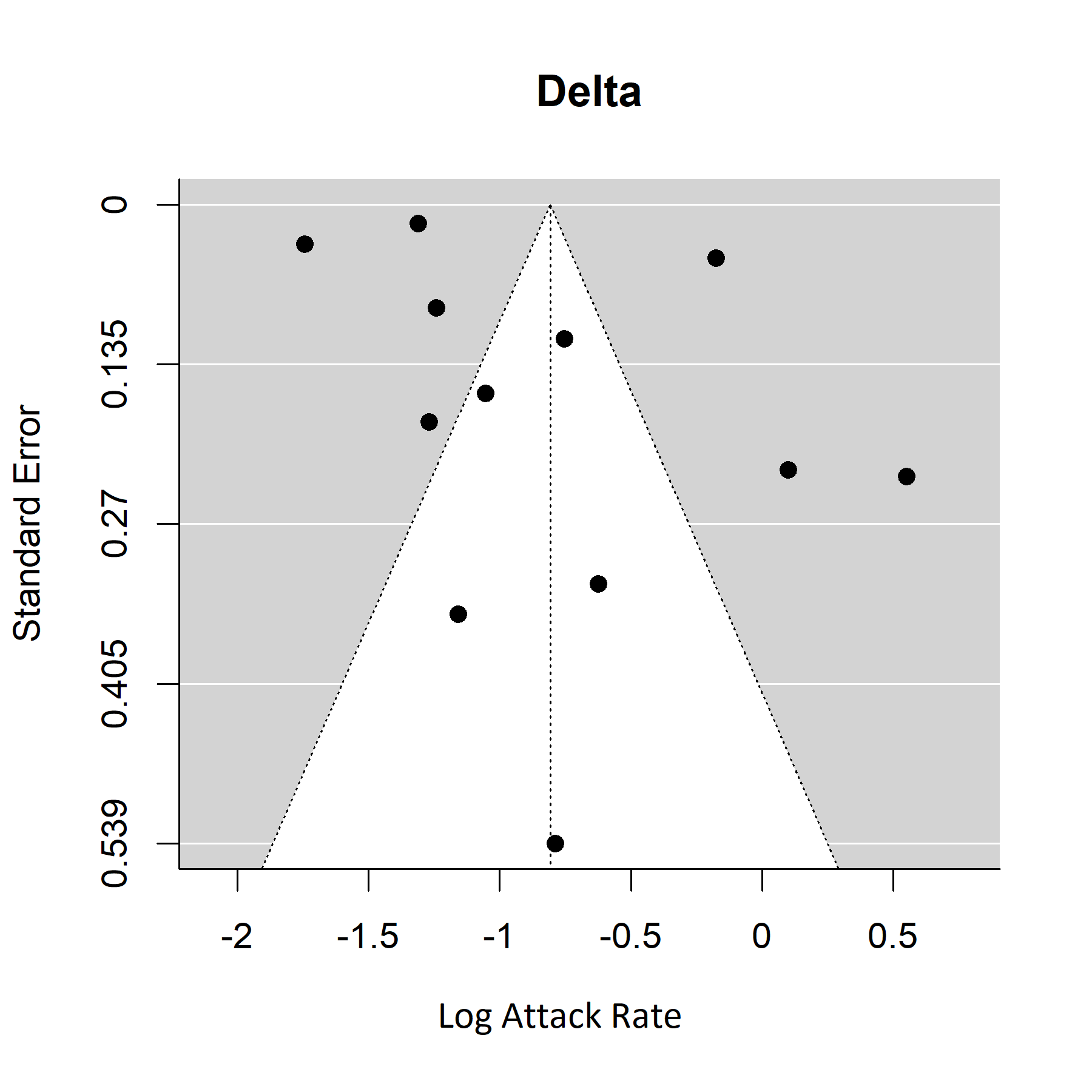

**S4 Figure. Household secondary attack rates for Delta (B.1.617.2) variant from unvaccinated index cases to unvaccinated household contacts.**

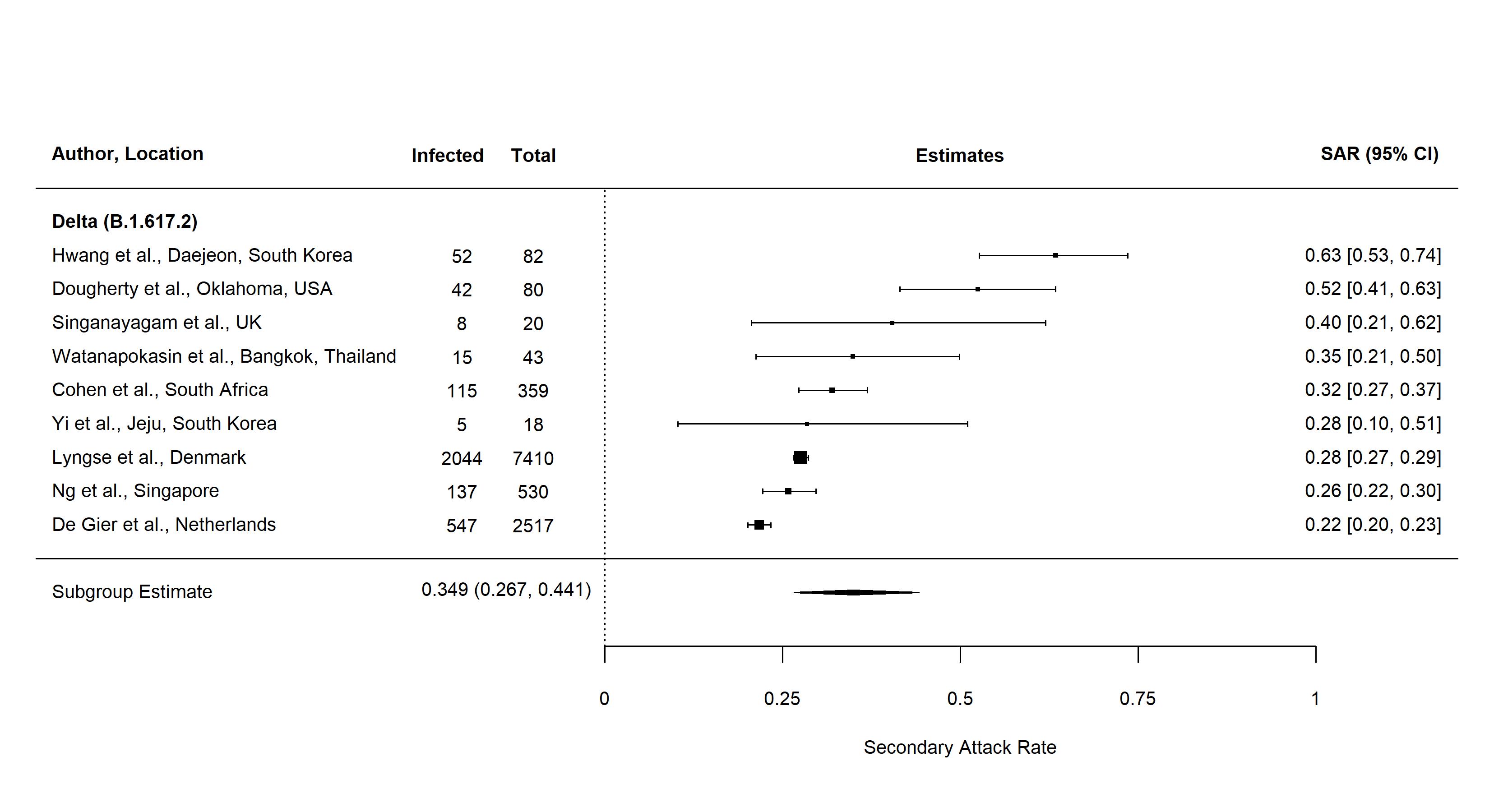

Three studies included did not disaggregate household secondary attack rates by vaccination status. Watanapokasin *et al*. reported that 83% of index cases were unvaccinated. Vaccination status was not provided for contacts. Dougherty *et al.* reported that 17 of 194 (8.8%) exposed individuals in the study were fully vaccinated. Cohen *et al.* reported that vaccine uptake was low in study sites reaching 5% fully vaccinated by the end of follow up.

**S5 Figure. Household secondary attack rates by index case vaccination status. Only unvaccinated contacts are included.**

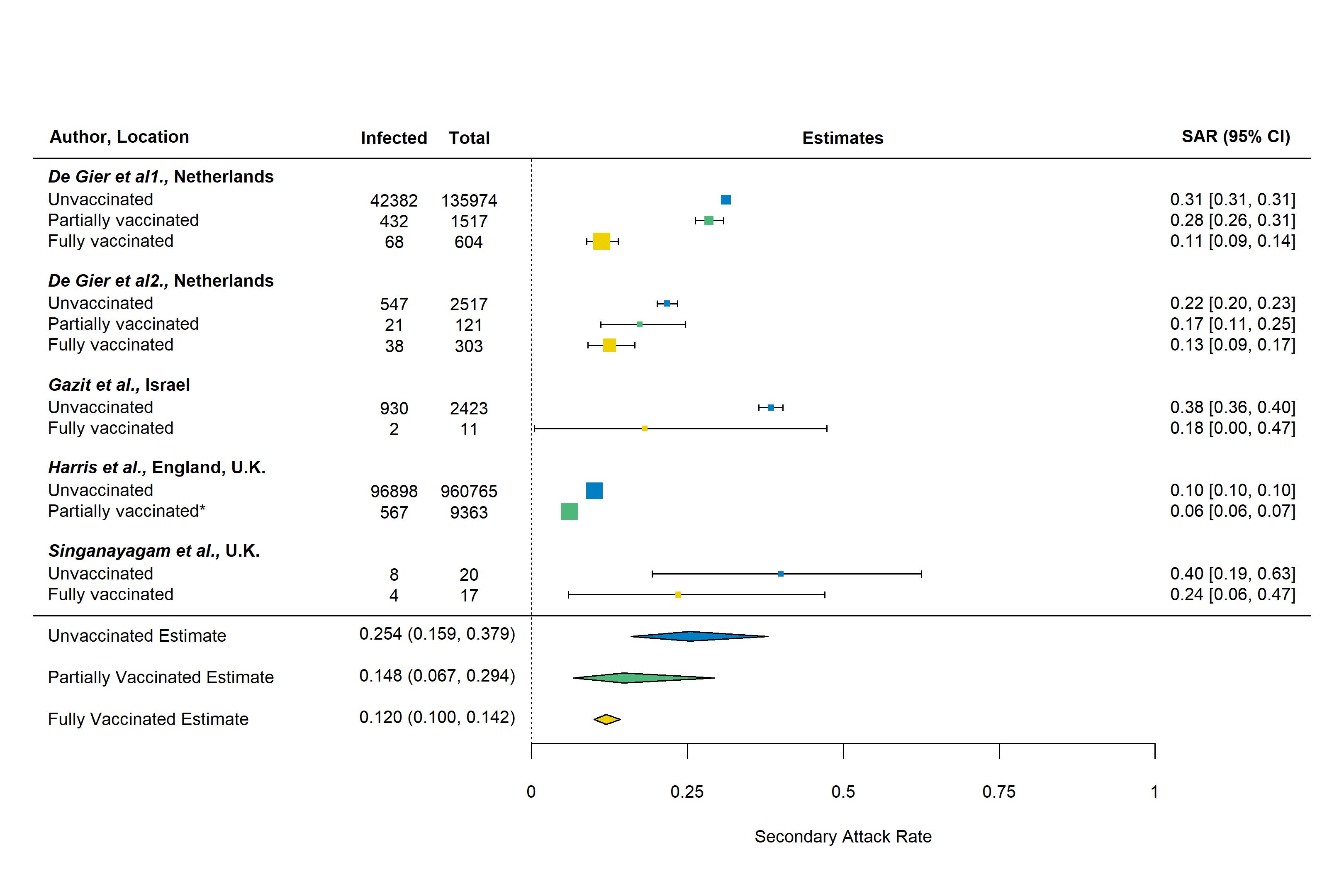

*For *Harris et al.*, most of the vaccinated index cases (93%) had received only the first dose of vaccine and secondary attack rates were not disaggregated by dose.

**S6 Figure. Household secondary attack rates by vaccination status of the index cases and contacts. The number of studies is provided.**

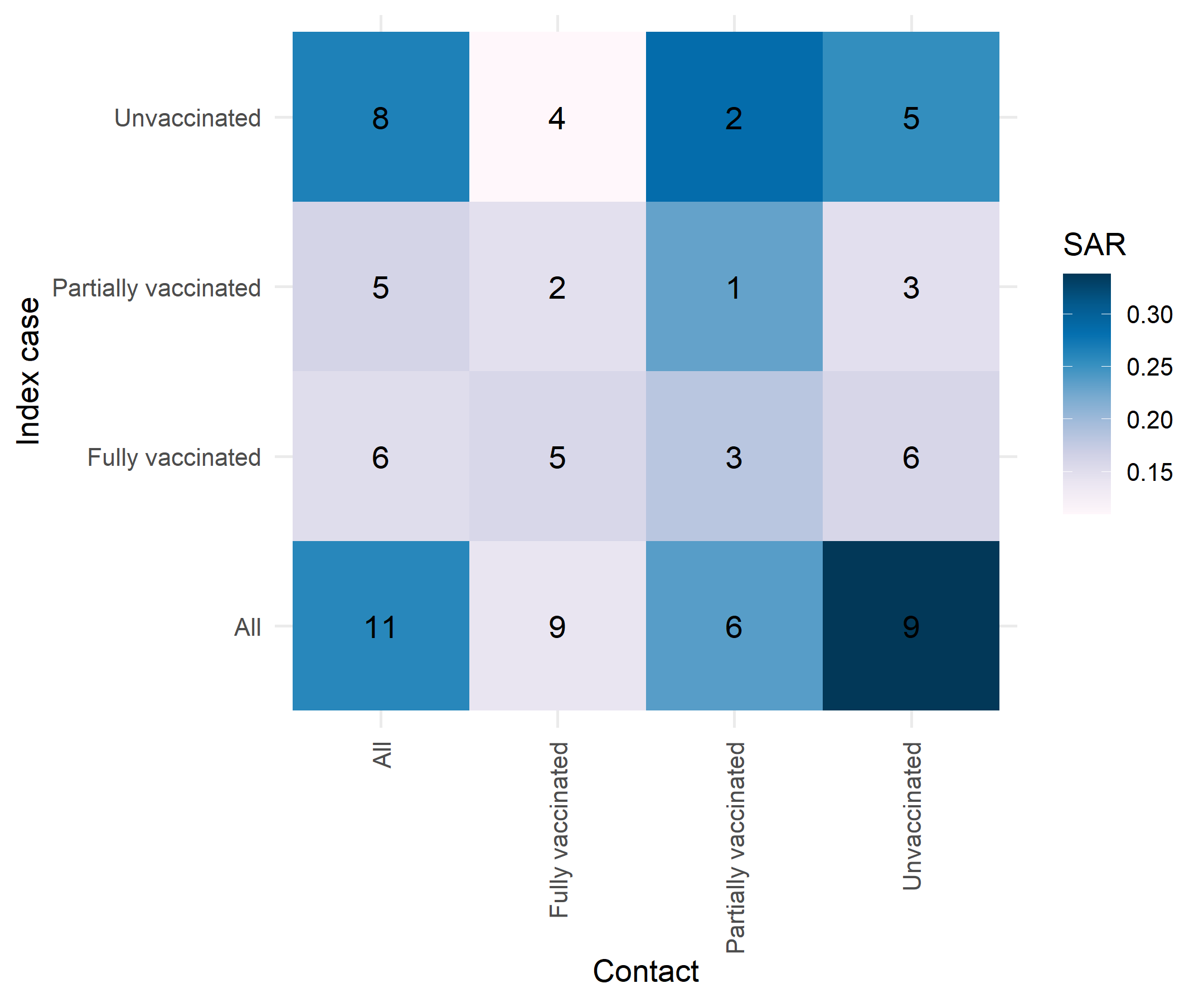

**S7 Figure. Household secondary attack rates by contact vaccination status. Only unvaccinated index cases are included.**

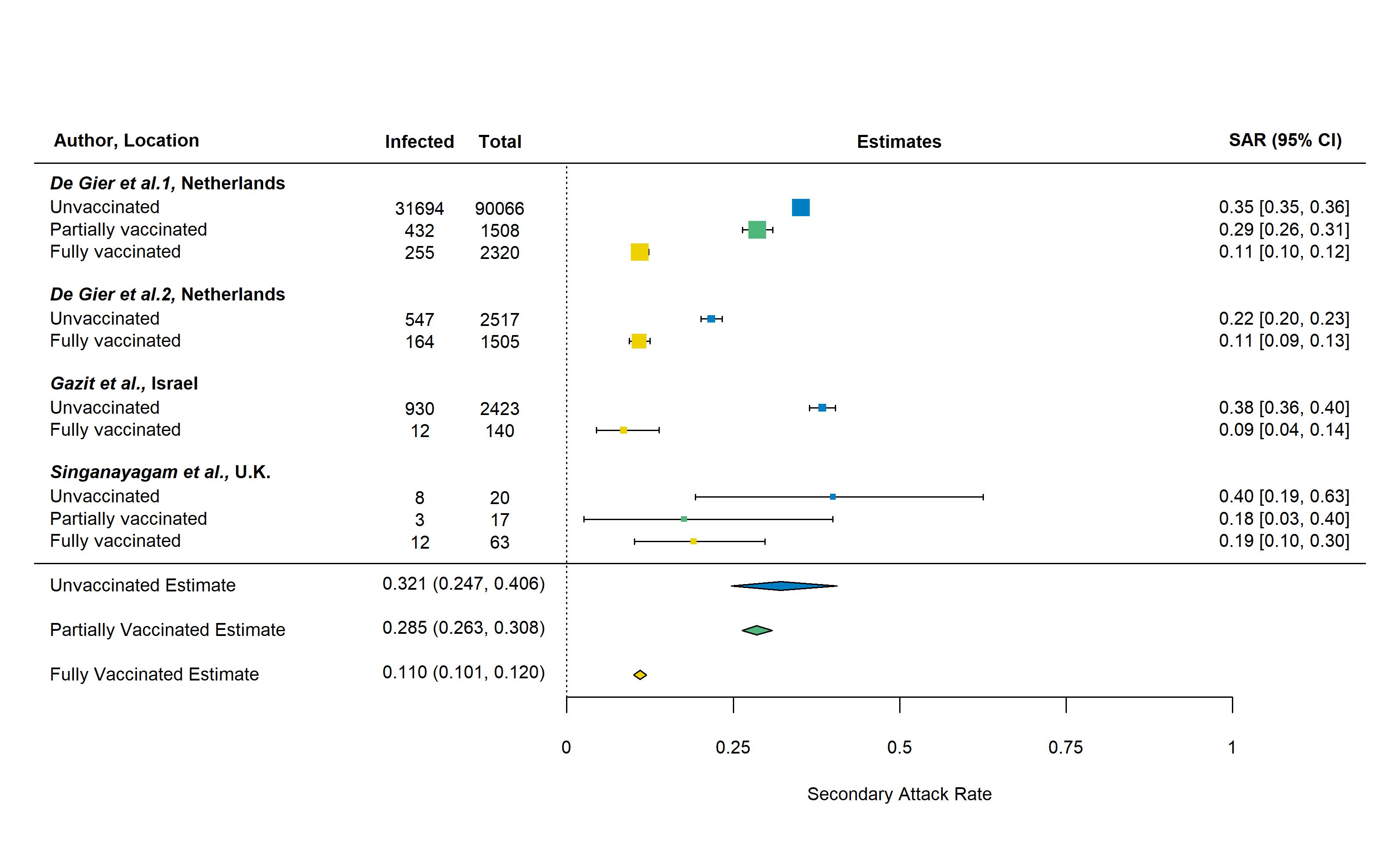

**S8 Figure. Household secondary attack rates for BNT162b2 vaccine by index case vaccination status. All contacts are included regardless of vaccination status.**

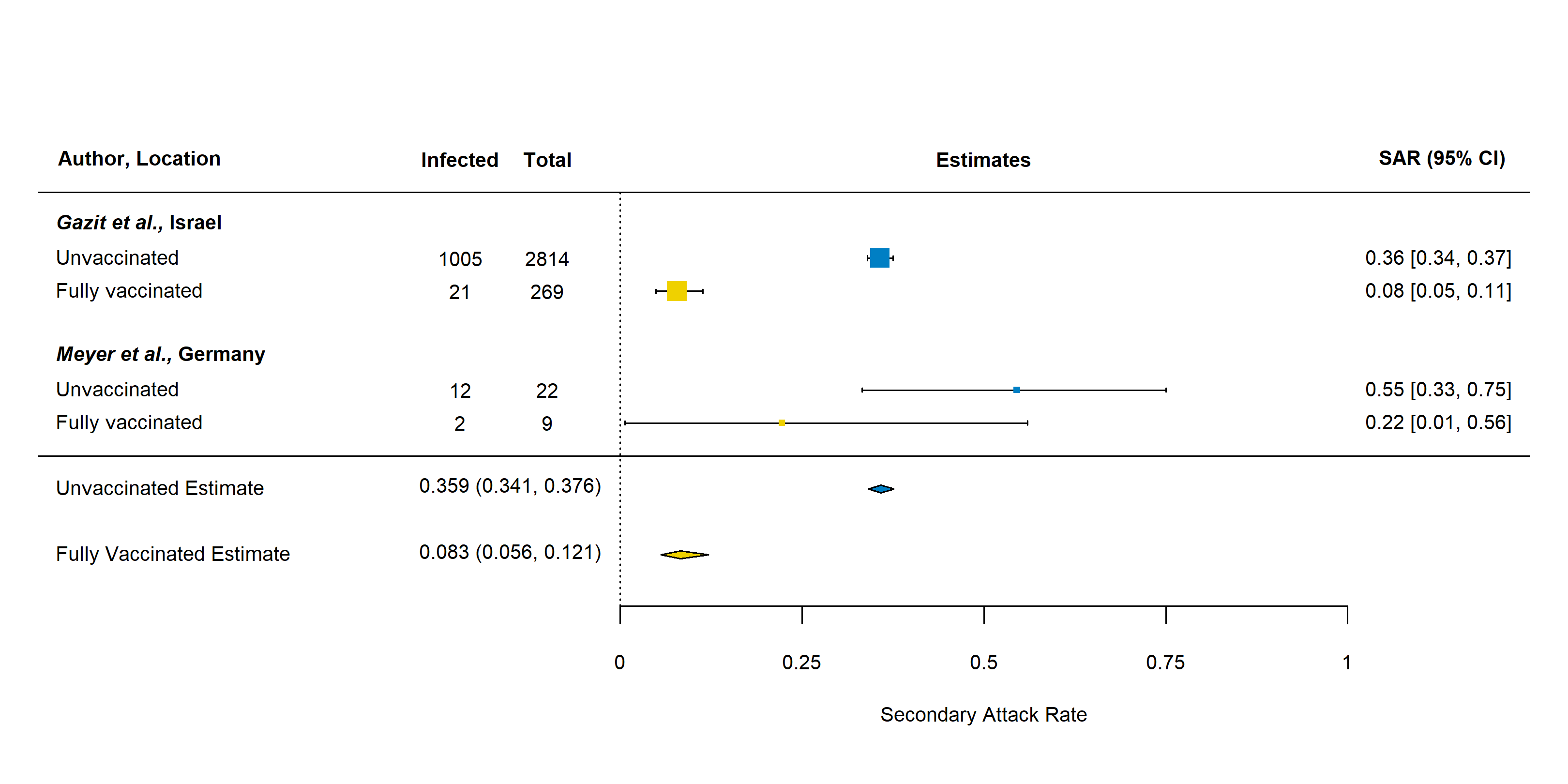

**S9 Figure. Household secondary attack rates by vaccine and contact vaccination status. All index cases are included regardless of vaccination status.**

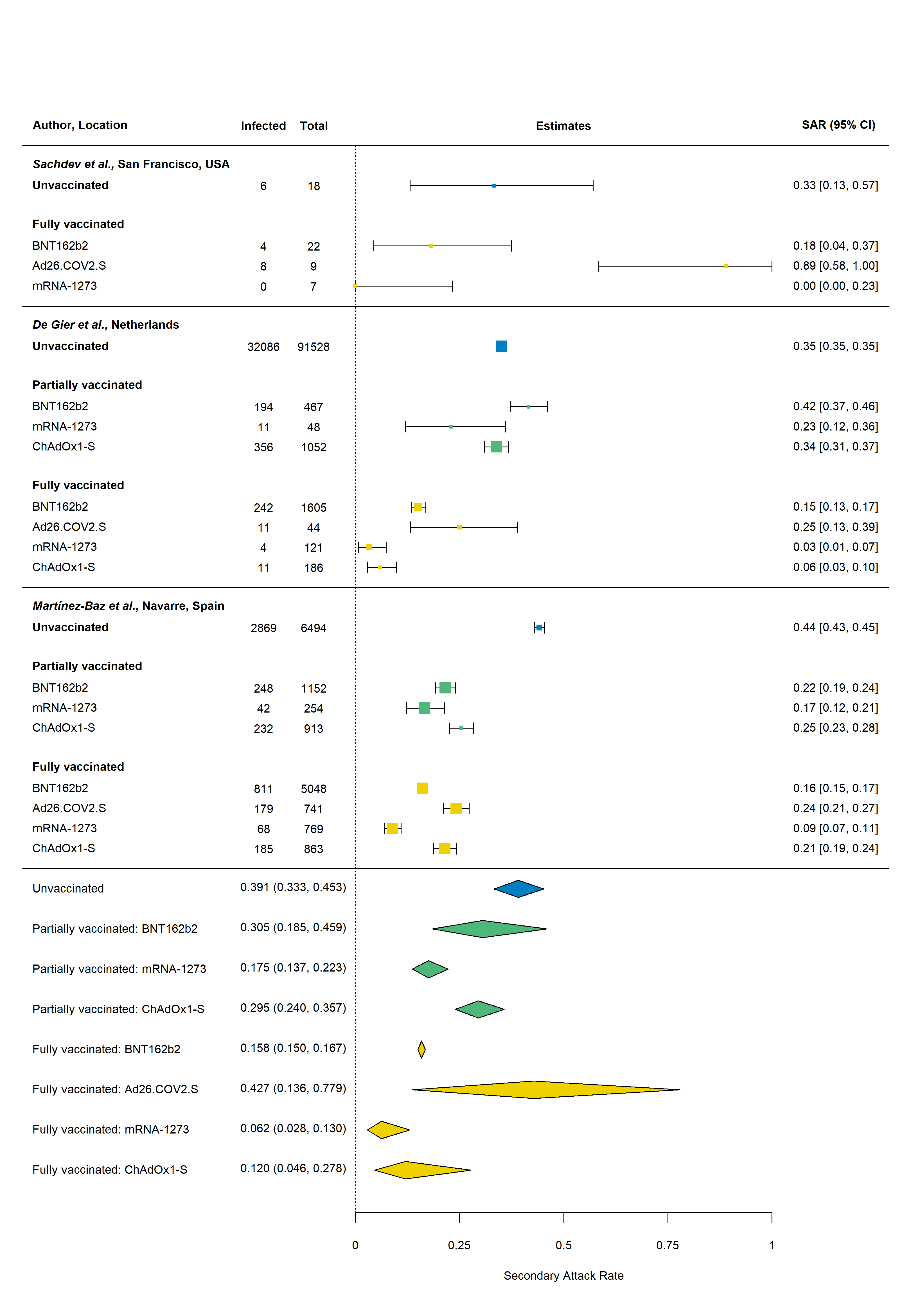

**S1 Table. Electronic databases and search strategy for household secondary attack rate of SARS-CoV-2.**

| **Database: PubMed: 1,287 retrieved articles** | | |
| --- | --- | --- |
| #1: "SARS-CoV-2" [All Fields] OR "COVID-19" [All Fields] OR "severe acute respiratory syndrome" [All Fields] OR "SARS" [All Fields] OR "SARS-CoV" [All Fields] OR "coronavirus" [All Fields] OR "variant" [All Fields] OR "vaccination" [All Fields] or “immunization” [All Fields] | #2: "secondary attack rate" [All Fields] OR "secondary infection rate" [All Fields] OR "household" [All Fields] OR "family contacts" [All Fields] OR "close contacts" [All Fields] OR "index case" [All Fields] OR "contact transmission" [All Fields] OR "contact attack rate" [All Fields] OR "family transmission" [All Fields] | "2021/06/18"[Date - Publication]: "3000"[Date - Publication] |
| **#1 AND #2 AND #3** | | |

**S2 Table. Description of studies identified from June 18, 2021 to January 7, 2022**

| **Authors** | **Location** | **Study period** | **No. index**  **cases** | **Index case**  **symptom status** | **Duration of follow-up (days)** | **Test used to diagnose contacts** | **Universal**  **testing or**  **only**  **symptomatic** | **Number of**  **tests per**  **contact** | **Household SAR (infected/total)** |
| --- | --- | --- | --- | --- | --- | --- | --- | --- | --- |
| *Afonso et al.^1^* | Goiânia, Brazil | June 15 – October 28, 2020 | 187 | Symptomatic and asymptomatic | 14 | RT-PCR | Universal | 1 | Untyped: 25.1% (67/267) |
| *Bistaraki et al.^2^* | Greece | October 1 – December 9, 2020 | – | Symptomatic and asymptomatic | – | – | – | – | Untyped: 18.3% (10,247/55,991) |
| *Burke et al.^3^* | USA | January 14 – April 4, 2020 | 69 | Symptomatic | 14 | Symptom-based diagnosis | Symptomatic | 0 | Untyped: 32% (62/193) |
| *Calvani et al.^4^* | Rome, Italy | October 16 –December 19, 2020 | 28 | Symptomatic and asymptomatic | – | – | Universal | 1 | Untyped: 30.6% (22/72) |
| *Cheng et al.^5^* | Hong Kong | April – May, 2021 | – | Symptomatic and asymptomatic | 14 or 21 | RT-PCR and whole genome sequencing | Universal | 4 | Beta: 28.0% (7/25) |
| *Chu et al.^6^* | Georgia, USA | July 17 –August 24, 2020 | 224 | Symptomatic and asymptomatic | – | Antigen test | Universal | At least 1 | Untyped: 12.2% (46/377) |
| *Clifford et al.^7^* | England | February 2, 2021 – September 10, 2021 | 195 | Symptomatic and asymptomatic | 14 | RT-PCR & genomic sequencing | Universal | 3 | Alpha: 49% (95% CI: 34-63%)  Delta: 81% (95% CI: 57%-96%)  Numerator/denominator provided for overall only: 40.6% (113/278) |
| *Cohen et al.^8^* | South Africa | July 16, 2020 – March 31, 2021 | 103 | Symptomatic and asymptomatic | 14 | RT-PCR | Universal | Multiple | Alpha: 57.1% (4/7)  Beta: 22.1% (81/366)  Delta: 32.0% (115/359)  Wild type: 10.5% (16/153) |
| *De Gier et al.^9^* | Netherlands | February 1 – May 27, 2021 | 85,210 | Symptomatic | 14 | RT-PCR, loop mediated isothermal amplification or antigen test | Universal | – | Untyped: 30.7% (43,735/142,540) |
| *De Gier et al.^10^* | Netherlands | August 9 – September 24, 2021 | 4,921 | Symptomatic | 10 | RT-PCR, loop mediated isothermal amplification or antigen test | Universal | – | Untyped: 14.9% (1,063/7,130) |
| *Dougherty et al.^11^* | Oklahoma, U.S.A. | April 15 – May 3, 2021 | 21 | Symptomatic and asymptomatic | 14 | Sequencing | Universal | 1 | Delta: 52.5% (42/80) |
| *Dub et al.^12^* | Helsinki, Finland | March 24, 2020 – June 17, 2020 | 39 | Symptomatic and asymptomatic | 28 | RT-PCR | Universal | 4 | Untyped: RT-PCR: 32.2% (28/87) |
| *Gazit et al.^13^* | Israel | December 20, 2020 –  March 17, 2021 | 2,827 | Symptomatic and asymptomatic | 10 | RT-PCR | Universal | 1 | Untyped: 34.1% (1,373/4,024) |
| *Ge et al.^14^* | Zhejiang Province, China | January 8 – July 30, 2020 | 370 | Symptomatic and asymptomatic | 14 | RT-PCR | Universal | Multiple | Untyped: 10.1% (260/2,565) |
| *Gorgels et al.^15^* | Netherlands | March – April, 2021 | 97 | Symptomatic and asymptomatic | 14 | RT-PCR or antigen test | Universal | At least 1 | Alpha: 39.8% (99/249) |
| *Hwang et al.^16^* | Daejeon, South Korea | June 22 –  July 31, 2021 | 23 | Symptomatic and asymptomatic | 14 | RT-PCR | Universal | 3 | Delta: 64.2% (52/82). |
| *Jagdale et al.^17^* | Pune City, India | April 1 – May 15, 2020 | 119 | Symptomatic and asymptomatic | 14 | RT-PCR | Universal | 1 | Untyped: 35% (198/565) |
| *Julin et al.^18^* | Oslo/Viken, Norway | May –June 2020, & September 2020 –end of April 2021 | 65 | Symptomatic | 14 | RT-PCR and next-generation sequencing | Universal | 8 | SAR of 77.8% reported for Alpha, however, footnote c of Table 2 indicates the SAR for Alpha includes, “households with at least one confirmed case among its household contacts.”  We therefore calculated the SAR as: (28-15)/(36-15)=61.9%, where 15 is the number of households with transmission.  SAR for All variants: (67-39)/(135-65)=40.0% |
| *Kang et al.^19^* | Guangdong, China | May 21 –June 18, 2021 | 73 | Symptomatic and asymptomatic | 14 | RT-PCR | Universal | Multiple | Delta: 22.0% (38/173) |
| *Karumanagoundar et al.^20^* | Tamil Nadu, India | March 14, 2020 – May 30, 2020 | 931 | Symptomatic and asymptomatic | 14 | RT-PCR | Universal | 1 | Untyped: 13.4% (464/3,474) |
| *Layan et al.^21^* | Israel | December 31 2020 –April 26 2021 | 212 | Symptomatic and asymptomatic | 10 | RT-PCR | Universal | 2 | Untyped: 41% (261/641) |
| *Li et al.^22^* | Hubei, China | January 23 – February 25, 2020 | 476 | Symptomatic | 14 | RT-PCR | Symptomatic | 1 | Untyped: 10.8% (110/1,020) |
| *Liu et al.^23^* | Los Angeles, USA | December 2020 – February 2021 | 15 | Symptomatic | 14 | RT-PCR | Universal | 5 | Untyped: 34.0% (17/50) |
| *Loss et al.^24^* | Germany | October 2020 – June 2021 | 24 | Symptomatic and asymptomatic | 12 | RT-PCR | Universal | Multiple | Untyped: 53.3% (24/45) |
| *Lyngse et al.^25^* | Denmark | December 9, 2021 – December 21, 2021 | Delta: 4629 unvaccinated;  Omicron: 368 unvaccinated | Symptomatic and asymptomatic | 7 | RT-PCR and antigen tests | Universal | 1 | Delta: 21.3% (4,923/23,156)  Omicron: 31.2% (1,474/4,718) |
| *Martinez et al.^26^* | Baltimore, USA | June 11, 2020 – May 20, 2021 | 277 | Symptomatic | 14 | RT-PCR | Universal | 1 | Untyped: 45.8% (292/638) |
| *Martínez-Baz et al.^27^* | Navarre, Spain | April – August 2021 | – | Symptomatic and asymptomatic | 10 | RT-PCR | Universal | 2 | Untyped: 28.5% (4,639/16,305) |
| *Meyer et al.^28^* | Germany | December 31, 2020 – April 26, 2021 | 14 | Symptomatic and asymptomatic | 14 | RT-PCR | Universal | 2 | Untyped: 45.2% (14/31) |
| *Miller et al.^29^* | England | March 30, 2020 – November 17, 2020 | 181 | Symptomatic | 14 | RT-PCR | Universal | 2 | Untyped: 21.1% (91/431) (PCR only) |
| *Ministry of Health NZ^30^* | New Zealand | August, 2021 | – | – | – | – | – | – | Delta: 45.6% (902/196) |
| *Montecucco et al.^31^* | Genoa, Italy | October 1, 2020 – March 31, 2021 | 6 | Symptomatic and asymptomatic | – | RT-PCR | Universal | 1 | Untyped: 33.3% (17/51) |
| *Musa et al.^32^* | Bosnia and Herzegovina | August 3 – December 23, 2020 | 360 | Symptomatic | 14 | RT-PCR or symptom-based diagnosis | – | – | Untyped: 15.9% (119/747) |
| *Ng et al.^33^* | Singapore | September 1, 2020 and May 31, 2021 | 301 | Symptomatic | 14 | RT-PCR & whole-genome sequencing | Universal | 2 | Delta: 22.4% (169/753)  Other: 13.3% (33/248) |
| *Ng et al.^34^* | Negeri Sembilan, Malaysia | February 1, 2020 –December 31, 2020 | 185 | Symptomatic and asymptomatic | 14 | RT-PCR | Universal | At least 1 | Untyped: 55.0% (466/848) |
| *Ogata et al.^35^* | Tsuchiura, Japan | August 2020 – February 2021 | 236 | Symptomatic and asymptomatic | – | RT-PCR | Universal | At least 1 | Untyped: 25.2% (125/496) |
| *Rajmohan et al.^36^* | Thrissur, India | January 1, 2021 – February 28, 2021 | 101 | Symptomatic and asymptomatic | 14 | RT-PCR or Rapid Antigen Test | Universal | 1 | Untyped: 40.7% (185/387) |
| *Ratovoson et al.^37^* | Madagascar | March 19 – July 30, 2020 | 33 | Symptomatic and asymptomatic | 21 | RT-PCR | Universal | 4 | Untyped: 38.8% (56/179) |
| *Remón-Berrade et al.^38^* | Navarre, Spain | March 2 – May 26, 2020 | 89 | Symptomatic | – | RT-PCR or Antigen Test | Universal | 1 | Untyped: 14.1% (46/326) |
| *Sachdev et al.^39^* | San Francisco, USA | January 29-July 2, 2021 | 105 | Symptomatic and asymptomatic | 14 | RT-PCR, loop-mediated amplification, or antigen Test | Universal | – | Overall: 28.2% (20/71)  Delta: 33.3% (5/16)  Alpha: 25.0% (2/8)  Gamma: 0% (0/3)  Beta: 20.0% (1/5)  Iota: 55.6% (5/9)  Kappa: 0% (0/2) |
| *Singanayagam et al.^40^* | U.K. | Sept 13, 2020, –Sept 15, 2021 | 138 | Symptomatic | 14-20 | RT-PCR & whole-genome sequencing | Universal | Daily | Delta: 25.9% (53/205) |
| *Sorioano-Arandes et al.^41^* | Catalonia, Spain | July 1, 2020 – October 31, 2020 | 80 | Symptomatic and asymptomatic | – | RT-PCR or antigen testing | Universal | – | Untyped: 64.8% (560/864) |
| *Tanaka et al.^42^* | Osaka Prefecture, Japan | December 1–20, 2020 & April 20, 2021 – May 3, 2021 | 307 | Symptomatic | 14 | RT-PCR | Universal | At least 1 | Pre-existing virus: 19.3% (56/290)  Alpha: 38.7% (48/124) |
| *ur Rehman et al.^43^* | Islamabad, Pakistan | March 21 – April 4, 2020 | – | Symptomatic | 14 | RT-PCR | Universal | 1 | Untyped: 36.8% (14/38) |
| *Watanapokasin et al.^44^* | Bangkok, Thailand | May 1 –June 30, 2021 | 30 | Symptomatic | 14 | RT-PCR | Universal | – | Alpha: 48.6% (17/35)  Delta: 34.9% (15/43). |
| *Yi et al.^45^* | Jeju, South Korea | August 3 – August 10, 2021 | 25 | Symptomatic and asymptomatic | 14 | RT-PCR | Universal | 1 | Delta: 23.9% (11/46) |

**S3 Table. References for studies included in analysis of SAR over time.**

| **Studies** | **References** |
| --- | --- |
| All 126 studies | ^1-126^ |
| 49 new studies (June 18, 2021 and January 7, 2022) | ^1-48,126^ |
| 77 studies from previous review^a^ (through June 17, 2021) | ^49-125^ |
| 27 studies with midpoints in 2021 | ^5,7-11,13,15,16,19,21,23-25,27,28,30,33,36,39,40,42,44,45,47,48,88^ |
| 62 studies with midpoints through April 2020 | ^3,14,17,20,22,43,53-58,60-62,64,66-69,72-76,79-81,83-87,89,90,92-97,99,100,102,104,105,107,109,111,113,115-125,127^ |
| ^a^5 studies were excluded that did not include laboratory-confirmed infections^128-132^ and 1 that included only asymptomatic index cases^127^ | |

**S4 Table. Risk of bias assessment for studies included in review of household transmissibility of SARS-CoV-2 using the same modified version of the Newcastle–Ottawa quality assessment scale for observational studies used by *Fung et al.^133^***

|  | **Selection** | | | **Comparability** | **Outcome** | | |  | |
| --- | --- | --- | --- | --- | --- | --- | --- | --- | --- |
| **Author** | **Representativeness of the index cases in region (2 points)^a^** | **Index case definition (1 point)^b^** | **Sample size (1 point)^c^** | **Household SAR disaggregated by index or contact covariates (1 point)^d^** | **Universal or symptom-based testing (1 point)^e^** | **Follow-up duration**  **(2 points)^f^** | **Number of tests per contact (1 point)^g^** | **Total points** | **Risk of bias^h^** |
| *Cheng et al.^5^* | + | + | 0 | + | + | ++ | + | 7 | Low |
| *Cohen et al.^8^* | ++ | + | + | + | + | ++ | + | 9 | Low |
| *De Gier et al.^9^* | ++ | + | + | + | + | ++ | 0 | 8 | Low |
| *De Gier et al.^10^* | ++ | + | + | + | + | ++ | 0 | 8 | Low |
| *Dougherty et al.^11^* | + | + | 0 | 0 | + | ++ | 0 | 5 | Moderate |
| *Gazit et al.^13^* | ++ | + | + | + | + | + | 0 | 7 | Low |
| *Gorgels et al.^15^* | + | + | 0 | + | + | ++ | + | 7 | Low |
| *Harris et al.^47^* | ++ | + | + | + | + | ++ | + | 9 | Low |
| *Hwang et al.^16^* | ++ | + | 0 | 0 | + | ++ | + | 7 | Low |
| *Julin et al.^18^* | + | + | 0 | + | + | ++ | + | 7 | Low |
| *Kang et al.^19^* | ++ | + | 0 | 0 | + | ++ | + | 7 | Low |
| *Layan et al.^21^* | + | + | + | + | + | + | + | 7 | Low |
| *Loenenbach et al.^88^* | + | + | 0 | + | 0 | ++ | 0 | 5 | Moderate |
| *Lyngse et al.^48^* | ++ | + | + | + | + | ++ | 0 | 8 | Low |
| *Lyngse et al.^25^* | ++ | + | + | + | + | + | 0 | 7 | Low |
| *Martínez-Baz et al.^27^* | ++ | + | + | + | + | + | + | 8 | Low |
| *Meyer et al.^28^* | 0 | + | 0 | + | + | ++ | + | 6 | Moderate |
| *Ministry of Health NZ^30^* | ++ | 0 | + | 0 | 0 | 0 | 0 | 3 | High |
| *Ng et al.^33^* | ++ | + | + | + | + | ++ | + | 9 | Low |
| *Sachdev et al.^39^* | + | + | 0 | + | + | ++ | 0 | 6 | Moderate |
| *Singanayagam et al.^40^* | + | + | 0 | + | + | ++ | + | 7 | Low |
| *Tanaka et al.^42^* | ++ | + | 0 | + | + | ++ | + | 8 | Low |
| *Watanapokasin et al.^44^* | 0 | + | 0 | + | + | ++ | 0 | 5 | Moderate |
| *Yi et al.^45^* | 0 | + | 0 | + | + | ++ | 0 | 5 | Moderate |

^a^ ++: Representative of COVID-19 cases in region; +: Somewhat representative; 0: Poorly described or not representative of cases in region

^b^ +: Index case identified by date of onset of symptoms and/or test dates; 0: First case not clearly defined

^c^ +: ≥300 contacts; 0: <300 contacts

^d^ +: Secondary attack rate disaggregated by ≥1 covariate; 0: Secondary attack rate not disaggregated by any covariates

^e^ +: Tested all contacts (both symptomatic and asymptomatic); 0: Only tested symptomatic contacts

^f^ ++: ≥14 days; +: 7 days; 0: <7 days or not specified

^g^ +: ≥2 tests; 0: 1 test or not described

^h^ High: ≤3 points; moderate: 4–6 points; low: ≥7 points

**S5 Table. Vaccination status definitions for studies that reported household secondary attack rates by vaccination status of index cases or contacts**

| **Study** | **Vaccination definitions** |
| --- | --- |
| *De Gier et al.^9^* | Partly vaccinated was defined as having received the first dose of a two-dose schedule at least 14 days before onset of symptoms. Fully vaccinated was defined as having completed a two-dose schedule at least 7 days or the one-dose Janssen schedule at least 14 days before symptom onset. |
| *De Gier et al.^10^* | Partly vaccinated was defined as having received the first dose of a 2‐dose schedule with a time since vaccination of at least 14 days. Fully vaccinated was defined as having completed a 2‐dose schedule with a time since vaccination of at least 14 days, or the 1‐dose Janssen schedule with a time since vaccination of at least 28 days. |
| *Harris et al.^47^* | Vaccinated index cases defined as having been vaccinated 21 days or more prior to testing positive for COVID-19 based on evidence of the time needed for the vaccine to provide a sufficient level of immunity. Non-vaccinated index cases were defined as not having received a vaccine prior to testing positive. Households where the index case received the vaccine less than 21 days before testing positive were excluded from this analysis.  Most of the vaccinated index patients (93%) had received only the first dose of vaccine. SAR not disaggregated by dose. |
| *Gazit et al.^13^* | Participants were classified into one of three vaccination-status groups at the time of the index case (the confirmed exposure): Unvaccinated; Recently Vaccinated Once, i.e. those vaccinated with the first vaccine dose within 0-7 days before the index infection, and Fully Vaccinated, i.e. those who were 7 or more days post the second dose by the time of the confirmed exposure. |
| *Layan et al.^21^* | Cases were considered vaccinated if their infection occurred >7 days after the 2nd dose. Similarly, household contacts were considered vaccinated if their exposure to the index case occurred >7 days after the 2nd dose |
| *Martínez-Baz et al.^27^* | A person was considered fully vaccinated ≥ 14 days after receiving one dose of Janssen or the second dose of other vaccines, and partially vaccinated ≥ 14 days after receiving only the first dose of Spikevax, Comirnaty or Vaxzevria. |
| *Meyer et al.^28^* | Not defined, but the two secondary cases found among household contacts of vaccinated index cases were diagnosed 25 days after the second vaccination. |
| *Ng et al.^33^* | Both index cases and close contacts were considered partially vaccinated if they had received one vaccine dose before the day the quarantine order was issued, or were within 14 days of the second dose on the day the quarantine order was issued. If more  than 14 days had elapsed after their second dose, they were taken to be fully vaccinated. |
| *Sachdev et al.^39^* | Partially vaccinated patients were defined as patients who received at least 1 dose of vaccine but were not fully vaccinated. Fully vaccinated patients were defined as patients who had received a second mRNA vaccine dose or a single-dose viral vector vaccine ≥14 days from symptom onset or collection of a positive specimen |
| *Singanayagam et al.^40^* | Participant defined as unvaccinated if they had not received a single dose of a COVID-19 vaccine at least 7 days before enrolment, partially vaccinated if they had received one vaccine dose at least 7 days before study enrolment, and fully vaccinated if they had received two doses of a COVID-19 vaccine at least 7 days before study enrolment. |

**References**

1.	Afonso ET, Marques SM, Costa LD, et al. Secondary household transmission of SARS‐CoV‐2 among children and adolescents: Clinical and epidemiological aspects. *Pediatric Pulmonology*. 2021;

2.	Bistaraki A, Roussos S, Tsiodras S, Sypsa V. Age-dependent effects on infectivity and susceptibility to SARS-CoV-2 infection: results from nationwide contact tracing data in Greece. *Infectious Diseases*. 2021:1-10.

3.	Burke RM, Calderwood L, Killerby ME, et al. Patterns of Virus Exposure and Presumed Household Transmission among Persons with Coronavirus Disease, United States, January–April 2020. *Emerging Infectious Diseases*. 2021;27(9):2323.

4.	Calvani M, Cantiello G, Cavani M, et al. Reasons for SARS-CoV-2 infection in children and their role in the transmission of infection according to age: a case-control study. *Ital J Pediatr*. Sep 27 2021;47(1):193. doi:10.1186/s13052-021-01141-1

5.	Cheng VC-C, Siu GK-H, Wong S-C, et al. Complementation of contact tracing by mass testing for successful containment of beta COVID-19 variant (SARS-CoV-2 VOC B. 1.351) epidemic in Hong Kong. *The Lancet Regional Health-Western Pacific*. 2021;17:100281.

6.	Chu VT, Yousaf AR, Chang K, et al. Household transmission of SARS-CoV-2 from children and adolescents. *New England Journal of Medicine*. 2021;385(10):954-956.

7.	Clifford S, Waight P, Hackman J, et al. Effectiveness of BNT162b2 and ChAdOx1 against SARS-CoV-2 household transmission: a prospective cohort study in England. *medRxiv*. 2021:2021.11.24.21266401. doi:10.1101/2021.11.24.21266401

8.	Cohen C, Kleynhans J, von Gottberg A, et al. SARS-CoV-2 incidence, transmission and reinfection in a rural and an urban setting: results of the PHIRST-C cohort study, South Africa, 2020-2021. *medRxiv*. 2021:2021.07.20.21260855. doi:10.1101/2021.07.20.21260855

9.	de Gier B, Andeweg S, Joosten R, et al. Vaccine effectiveness against SARS-CoV-2 transmission and infections among household and other close contacts of confirmed cases, the Netherlands, February to May 2021. *Eurosurveillance*. 2021;26(31):2100640.

10.	de Gier B, Andeweg S, Backer JA, et al. Vaccine effectiveness against SARS-CoV-2 transmission to household contacts during dominance of Delta variant (B.1.617.2), August-September 2021, the Netherlands. *medRxiv*. 2021:2021.10.14.21264959. doi:10.1101/2021.10.14.21264959

11.	Dougherty K, Mannell M, Naqvi O, Matson D, Stone J. SARS-CoV-2 B. 1.617. 2 (Delta) variant COVID-19 outbreak associated with a gymnastics facility—Oklahoma, April–May 2021. *Morbidity and Mortality Weekly Report*. 2021;70(28):1004.

12.	Dub T, Nohynek H, Hagberg L, et al. High secondary attack rate and persistence of SARS-CoV-2 antibodies in household transmission study participants, Finland 2020. 2021;

13.	Gazit S, Mizrahi B, Kalkstein N, et al. BNT162b2 mRNA Vaccine Effectiveness Given Confirmed Exposure: Analysis of Household Members of COVID-19 Patients. *Clinical Infectious Diseases*. 2021;doi:10.1093/cid/ciab973

14.	Ge Y, Martinez L, Sun S, et al. COVID-19 transmission dynamics among close contacts of index patients with COVID-19: a population-based cohort study in Zhejiang province, China. *JAMA Internal Medicine*. 2021;181(10):1343-1350.

15.	Gorgels K, Alphen L, Hackert BvdVV, et al. Increased Transmissibility of SARS-CoV-2 Alpha Variant (B.1.1.7) in Children: Three Large Primary School Outbreaks Revealed by Whole Genome Sequencing in the Netherlands. *Research Square*. 2021/12/10 2021;doi:10.21203/rs.3.rs-1107495/v1

16.	Hwang H, Lim J-S, Song S-A, et al. Transmission dynamics of the Delta variant of SARS-CoV-2 infections in South Korea. *The Journal of Infectious Diseases*. 2021;doi:10.1093/infdis/jiab586

17.	Jagdale GR, Parande MA, Borle P, et al. Secondary Attack Rate among the Contacts of COVID-19 Patients at the Beginning of the Pandemic in Pune City of Western Maharashtra, India. *Journal of Communicable Diseases (E-ISSN: 2581-351X & P-ISSN: 0019-5138)*. 2021;53(3):51-59.

18.	Julin CH, Robertson AH, Hungnes O, et al. Household Transmission of SARS-CoV-2: A Prospective Longitudinal Study Showing Higher Viral Load and Increased Transmissibility of the Alpha Variant Compared to Previous Strains. *Microorganisms*. 2021;9(11):2371.

19.	Kang M, Xin H, Yuan J, et al. Transmission dynamics and epidemiological characteristics of Delta variant infections in China. *Medrxiv*. 2021;

20.	Karumanagoundar K, Raju M, Ponnaiah M, et al. Secondary attack rate of COVID-19 among contacts and risk factors, Tamil Nadu, March–May 2020: a retrospective cohort study. *BMJ open*. 2021;11(11):e051491.

21.	Layan M, Gilboa M, Gonen T, et al. Impact of BNT162b2 vaccination and isolation on SARS-CoV-2 transmission in Israeli households: an observational study. *MedRxiv*. 2021;

22.	Li Y, Liu J, Yang Z, et al. Transmission of Severe Acute Respiratory Syndrome Coronavirus 2 to Close Contacts, China, January–February 2020. *Emerging Infectious Diseases*. 2021;27(9):2288.

23.	Liu PY, Gragnani CM, Timmerman J, et al. Pediatric Household Transmission of Severe Acute Respiratory Coronavirus-2 Infection—Los Angeles County, December 2020 to February 2021. *The Pediatric Infectious Disease Journal*. 2021;40(10):e379.

24.	Loss J, Wurm J, Varnaccia G, et al. Transmission of SARS-CoV-2 among children and staff in German daycare centers: results from the COALA study. *medRxiv*. 2021:2021.12.21.21268157. doi:10.1101/2021.12.21.21268157

25.	Lyngse FP, Mortensen LH, Denwood MJ, et al. SARS-CoV-2 Omicron VOC Transmission in Danish Households. *medRxiv*. 2021:2021.12.27.21268278. doi:10.1101/2021.12.27.21268278

26.	Martinez DA, Klein EY, Parent C, et al. Latino Household Transmission of Severe Acute Respiratory Syndrome Coronavirus 2. *Clinical Infectious Diseases*. 2021;

27.	Martínez-Baz I, Trobajo-Sanmartín C, Miqueleiz A, et al. Product-specific COVID-19 vaccine effectiveness against secondary infection in close contacts, Navarre, Spain, April to August 2021. *Eurosurveillance*. 2021;26(39):2100894.

28.	Meyer ED, Sandfort M, Bender J, et al. Two doses of the mRNA BNT162b2 vaccine reduce severe outcomes, viral load and secondary attack rate: evidence from a SARS-CoV-2 Alpha outbreak in a nursing home in Germany, January-March 2021. *medRxiv*. 2021:2021.09.13.21262519. doi:10.1101/2021.09.13.21262519

29.	Miller E, Waight PA, Andrews NJ, et al. Transmission of SARS-CoV-2 in the household setting: A prospective cohort study in children and adults in England. *Journal of Infection*. 2021;83(4):483-489.

30.	Ministry of Health NZ. COVID-19 Variants Update. December 3, 2021. <https://www.health.govt.nz/system/files/documents/pages/22-november-2021-variants-update-summary.pdf>

31.	Montecucco A, Dini G, Rahmani A, et al. Investigating SARS-CoV-2 transmission among co-workers in a University of Northern Italy during COVID-19 pandemic: an observational study. *La Medicina del Lavoro | Work, Environment and Health*. 12/23 2021;112(6):429-435. doi:10.23749/mdl.v112i6.12527

32.	Musa S, Kissling E, Valenciano M, et al. Household transmission of SARS-CoV-2: a prospective observational study in Bosnia and Herzegovina, August–December 2020. *International Journal of Infectious Diseases*. 2021;112:352-361.

33.	Ng OT, Koh V, Chiew CJ, et al. Impact of Delta Variant and Vaccination on SARS-CoV-2 Secondary Attack Rate Among Household Close Contacts. *The Lancet Regional Health-Western Pacific*. 2021;17:100299.

34.	Ng DCE, Tan KK, Chin L, et al. Risk factors associated with household transmission of SARS‐CoV‐2 in Negeri Sembilan, Malaysia. *Journal of paediatrics and child health*. 2021;

35.	Ogata T, Irie F, Ogawa E, et al. Secondary Attack Rate among Non-Spousal Household Contacts of Coronavirus Disease 2019 in Tsuchiura, Japan, August 2020–February 2021. *International Journal of Environmental Research and Public Health*. 2021;18(17):8921.

36.	Rajmohan P, Jose P, Thodi JBA, et al. Dynamics of transmission of COVID-19 cases and household contacts: A prospective cohort study. *Journal of Acute Disease*. 2021;10(4):162.

37.	Ratovoson R, Razafimahatratra R, Randriamanantsoa L, et al. Household transmission of COVID-19 among the earliest cases in Antananarivo, Madagascar. *Influenza Other Respir Viruses*. Aug 10 2021;doi:10.1111/irv.12896

38.	Remón-Berrade M, Guillen-Aguinaga S, Sarrate-Adot I, et al. Risk of Secondary Household Transmission of COVID-19 from Health Care Workers in a Hospital in Spain. *Epidemiologia*. 2022;3(1):1-10.

39.	Sachdev DD, Chew Ng R, Sankaran M, et al. Contact tracing outcomes among household contacts of fully vaccinated COVID-19 patients - San Francisco, California, January 29-July 2, 2021. *Clin Infect Dis*. Dec 20 2021;doi:10.1093/cid/ciab1042

40.	Singanayagam A, Hakki S, Dunning J, et al. Community transmission and viral load kinetics of the SARS-CoV-2 delta (B. 1.617. 2) variant in vaccinated and unvaccinated individuals in the UK: a prospective, longitudinal, cohort study. *The Lancet Infectious Diseases*. 2021;

41.	Soriano-Arandes A, Gatell A, Serrano P, et al. Household Severe Acute Respiratory Syndrome Coronavirus 2 Transmission and Children: A Network Prospective Study. *Clin Infect Dis*. Sep 15 2021;73(6):e1261-e1269. doi:10.1093/cid/ciab228

42.	Tanaka H, Hirayama A, Nagai H, et al. Increased transmissibility of the SARS-CoV-2 alpha variant in a japanese population. *International Journal of Environmental Research and Public Health*. 2021;18(15):7752.

43.	ur Rehman S, Qaisrani M, Abbasi S, et al. COVID-19 outbreak in Islamabad resulting from a travel-associated primary case: A case series. *Global Biosecurity*. 2021;3(1)

44.	Watanapokasin N, Siripongboonsitti T, Ungtrakul T, et al. Transmissibility of SARS-CoV-2 variants as a secondary attack in Thai households: a retrospective study. *IJID Regions*. 2021;1:1-2.

45.	Yi S, Kim JM, Choe YJ, et al. SARS-CoV-2 Delta Variant Breakthrough Infection and Onward Secondary Transmission in Household. *J Korean Med Sci*. 1/ 2022;37(1):0.

46.	Cerami C, Popkin-Hall ZR, Rapp T, et al. Household transmission of SARS-CoV-2 in the United States: living density, viral load, and disproportionate impact on communities of color. *Clin Infect Dis*. Aug 12 2021;doi:10.1093/cid/ciab701

47.	Harris RJ, Hall JA, Zaidi A, Andrews NJ, Dunbar JK, Dabrera G. Effect of Vaccination on Household Transmission of SARS-CoV-2 in England. *New England Journal of Medicine*. 2021;

48.	Lyngse FP, Mølbak K, Skov RL, et al. Increased transmissibility of SARS-CoV-2 lineage B.1.1.7 by age and viral load. *Nature Communications*. 2021/12/13 2021;12(1):7251. doi:10.1038/s41467-021-27202-x

49.	Adamik B, Bawiec M, Bezborodov V, et al. Bounds on the total number of SARS-CoV-2 infections: The link between severeness rate, household attack rate and the number of undetected cases. 2020;

50.	Akaishi T, Kushimoto S, Katori Y, et al. COVID-19 transmission in group living environments and households. *Scientific Reports*. 2021/06/02 2021;11(1):11616. doi:10.1038/s41598-021-91220-4

51.	Areekal B, Vijayan S, Suseela MS, et al. Risk Factors, Epidemiological and Clinical Outcome of Close Contacts of COVID-19 Cases in a Tertiary Hospital in Southern India. *Journal of Clinical & Diagnostic Research*. 2021;15(3)

52.	Awang H, Yaacob EL, Syed Aluawi SN, et al. A case–control study of determinants for COVID-19 infection based on contact tracing in Dungun district, Terengganu state of Malaysia. *Infectious Diseases*. 2021;53(3):222-225.

53.	Bae S, Kim H, Jung T-Y, et al. Epidemiological Characteristics of COVID-19 Outbreak at Fitness Centers in Cheonan, Korea. *J Korean Med Sci*. 8/ 2020;35(31)

54.	Bender JK, Brandl M, Höhle M, Buchholz U, Zeitlmann N. Analysis of asymptomatic and presymptomatic transmission in SARS-CoV-2 outbreak, Germany, 2020. *Emerging infectious diseases*. 2021;27(4):1159.

55.	Bi Q, Wu Y, Mei S, et al. Epidemiology and transmission of COVID-19 in 391 cases and 1286 of their close contacts in Shenzhen, China: a retrospective cohort study. *The Lancet Infectious Diseases*. 2020;

56.	Böhmer MM, Buchholz U, Corman VM, et al. Investigation of a COVID-19 outbreak in Germany resulting from a single travel-associated primary case: a case series. *The Lancet Infectious Diseases*. 2020;

57.	Boscolo-Rizzo P, Borsetto D, Spinato G, et al. New onset of loss of smell or taste in household contacts of home-isolated SARS-CoV-2-positive subjects. *European Archives of Oto-rhino-laryngology*. May 24 2020:1-4. doi:10.1007/s00405-020-06066-9

58.	Burke RM. Active monitoring of persons exposed to patients with confirmed COVID-19—United States, January–February 2020. *MMWR Morbidity and mortality weekly report*. 2020;69

59.	Charbonnier L, Rouprêt-Serzec J, Caseris M, et al. Contribution of Serological Rapid Diagnostic Tests to the Strategy of Contact Tracing in Households Following SARS-CoV-2 Infection Diagnosis in Children. Brief Research Report. *Frontiers in Pediatrics*. 2021-May-10 2021;9(217)doi:10.3389/fped.2021.638502

60.	Chaw L, Koh WC, Jamaludin SA, Naing L, Alikhan MF, Wong J. Analysis of SARS-CoV-2 transmission in different settings, Brunei. *Emerging infectious diseases*. 2020;26(11):2598.

61.	Chen Y, Wang AH, Yi B, et al. [Epidemiological characteristics of infection in COVID-19 close contacts in Ningbo city]. *Zhonghua Liu Xing Bing Xue Za Zhi*. May 10 2020;41(5):667-671. doi:10.3760/cma.j.cn112338-20200304-00251

62.	Cheng HY, Jian SW, Liu DP, Ng TC, Huang WT, Lin HH. Contact Tracing Assessment of COVID-19 Transmission Dynamics in Taiwan and Risk at Different Exposure Periods Before and After Symptom Onset. *JAMA Intern Med*. May 1 2020;doi:10.1001/jamainternmed.2020.2020

63.	Dattner I, Goldberg Y, Katriel G, et al. The role of children in the spread of COVID-19: Using household data from Bnei Brak, Israel, to estimate the relative susceptibility and infectivity of children. *PLoS computational biology*. 2021;17(2):e1008559.

64.	Dawson P, Rabold EM, Laws RL, et al. Loss of Taste and Smell as Distinguishing Symptoms of COVID-19. *Clin Infect Dis*. Jun 21 2020;doi:10.1093/cid/ciaa799

65.	Demko ZO, Antar AAR, Blair PW, et al. Clustering of SARS-CoV-2 infections in households of patients diagnosed in the outpatient setting in Baltimore, MD. *Open Forum Infectious Diseases*. 2021;doi:10.1093/ofid/ofab121

66.	Dong XC, Li JM, Bai JY, et al. [Epidemiological characteristics of confirmed COVID-19 cases in Tianjin]. *Zhonghua Liu Xing Bing Xue Za Zhi*. May 10 2020;41(5):638-641. doi:10.3760/cma.j.cn112338-20200221-00146

67.	Doung-ngern P, Suphanchaimat R, Panjagampatthana A, et al. Case-Control Study of Use of Personal Protective Measures and Risk for Severe Acute Respiratory Syndrome Coronavirus 2 Infection, Thailand. *Emerging Infectious Diseases*. 2020;26(11)

68.	Draper AD, Dempsey KE, Boyd RH, et al. The first 2 months of COVID-19 contact tracing in the Northern Territory of Australia, March-April 2020. *Communicable Diseases Intelligence*. Jul 2 2020;44doi:10.33321/cdi.2020.44.53

69.	Fateh-Moghadam P, Battisti L, Molinaro S, et al. Contact tracing during Phase I of the COVID-19 pandemic in the Province of Trento, Italy: key findings and recommendations. *medRxiv*. 2020;

70.	Gomaa MR, El Rifay AS, Shehata M, et al. Incidence, household transmission, and neutralizing antibody seroprevalence of Coronavirus Disease 2019 in Egypt: Results of a community-based cohort. *PLOS Pathogens*. 2021;17(3):e1009413. doi:10.1371/journal.ppat.1009413

71.	Grijalva CG, Rolfes MA, Zhu Y, et al. Transmission of SARS-COV-2 infections in households—Tennessee and Wisconsin, April–September 2020. *Morbidity and Mortality Weekly Report*. 2020;69(44):1631.

72.	Han T. Outbreak investigation: transmission of COVID-19 starting from a spa facility in a local community in Korea. *Epidemiol Health*. 2020;0(0):e2020056-0. doi:10.4178/epih.e2020056

73.	Hsu C-Y, Wang J-T, Huang K-C, Chiao-Hsin Fan A, Yeh Y-P, Li-Sheng Chen S. Household Transmission but without the Community-acquired Outbreak of COVID-19 in Taiwan. *Journal of the Formosan Medical Association*. 2021/05/03/ 2021;doi:<https://doi.org/10.1016/j.jfma.2021.04.021>

74.	Hu P, Ma M, Jing Q, et al. Retrospective study identifies infection related risk factors in close contacts during COVID-19 epidemic. *International Journal of Infectious Diseases*. 2021;103:395-401.

75.	Hu S, Wang W, Wang Y, et al. Infectivity, susceptibility, and risk factors associated with SARS-CoV-2 transmission under intensive contact tracing in Hunan, China. *Nature Communications*. 2021/03/09 2021;12(1):1533. doi:10.1038/s41467-021-21710-6

76.	Hua CZ, Miao ZP, Zheng JS, et al. Epidemiological features and viral shedding in children with SARS-CoV-2 infection. *Journal of Medical Virology*. Jun 15 2020;doi:10.1002/jmv.26180

77.	Islam SS, Noman ASM. Transmission Dynamics and Contact Tracing Assessment of COVID-19 in Chattogram, Bangladesh and Potential Risk of Close Contacts at Different Exposure Settings. *Bangladesh and Potential Risk of Close Contacts at Different Exposure Settings*.

78.	Jashaninejad R, Doosti-Irani A, Karami M, Keramat F, Mirzaei M. Transmission of COVID-19 and its Determinants among Close Contacts of COVID-19 Patients Running title. *Journal of Research in Health Sciences*. 2021;

79.	Jing Q-L, Liu M-J, Yuan J, et al. Household secondary attack rate of COVID-19 and associated determinants in Guangzhou, China: a retrospective cohort study. *The Lancet Infectious Diseases*. 2020;

80.	Kim J, Choe YJ, Lee J, et al. Role of children in household transmission of COVID-19. *Archives of Disease in Childhood*. 2020:archdischild-2020-319910. doi:10.1136/archdischild-2020-319910

81.	Covid-19 National Emergency Response Center Epidemiology Case Management Team Korea Centers for Disease Control Prevention. Coronavirus Disease-19: Summary of 2,370 Contact Investigations of the First 30 Cases in the Republic of Korea. *Osong Public Health Res Perspect*. 2020;11(2):81-84. doi:10.24171/j.phrp.2020.11.2.04

82.	Koureas M, Speletas M, Bogogiannidou Z, et al. Transmission Dynamics of SARS-CoV-2 during an Outbreak in a Roma Community in Thessaly, Greece—Control Measures and Lessons Learned. *International Journal of Environmental Research and Public Health*. 2021;18(6):2878.

83.	Kuba Y, Shingaki A, Nidaira M, et al. The characteristics of household transmission during COVID-19 outbreak in Okinawa, Japan from February to May 2020. *Japanese Journal of Infectious Diseases*. 2021:JJID. 2020.943.

84.	Laxminarayan R, Wahl B, Dudala SR, et al. Epidemiology and transmission dynamics of COVID-19 in two Indian states. *Science*. Sep 30 2020;

85.	Lewis NM, Chu VT, Ye D, et al. Household transmission of SARS-CoV-2 in the United States. *Clinical Infectious Diseases*. 2020;

86.	Li F, Li Y-Y, Liu M-J, et al. Household transmission of SARS-CoV-2 and risk factors for susceptibility and infectivity in Wuhan: a retrospective observational study. *The Lancet Infectious Diseases*. 2021;

87.	Liu T, Liang W, Zhong H, et al. Risk factors associated with COVID-19 infection: a retrospective cohort study based on contacts tracing. *Emerging Microbes & Infections*. Jul 1 2020:1-31. doi:10.1080/22221751.2020.1787799

88.	Loenenbach A, Markus I, Lehfeld A-S, et al. SARS-CoV-2 variant B. 1.1. 7 susceptibility and infectiousness of children and adults deduced from investigations of childcare centre outbreaks, Germany, 2021. *Eurosurveillance*. 2021;26(21):2100433.

89.	Lopez Bernal J, Panagiotopoulos N, Byers C, et al. Transmission dynamics of COVID-19 in household and community settings in the United Kingdom. *medRxiv*. 2020:2020.08.19.20177188. doi:10.1101/2020.08.19.20177188

90.	Luo L, Liu D, Liao X, et al. Contact Settings and Risk for Transmission in 3410 Close Contacts of Patients With COVID-19 in Guangzhou, China: A Prospective Cohort Study. *Annals of internal medicine*. Aug 13 2020;doi:10.7326/m20-2671

91.	Lyngse FP, Kirkeby CT, Halasa T, et al. COVID-19 Transmission Within Danish Households: A Nationwide Study from Lockdown to Reopening. *medRxiv*. 2020;

92.	Malheiro R, Figueiredo AL, Magalhães JP, et al. Effectiveness of contact tracing and quarantine on reducing COVID-19 transmission: a retrospective cohort study. *Public Health*. 2020;

93.	Metlay JP, Haas JS, Soltoff AE, Armstrong KA. Household Transmission of SARS-CoV-2. *JAMA Network Open*. 2021;4(2):e210304-e210304.

94.	Miyahara R, Tsuchiya N, Yasuda I, et al. Familial Clusters of Coronavirus Disease in 10 Prefectures, Japan, February− May 2020. *Emerging infectious diseases*. 2021;27(3):915.

95.	Ng OT, Marimuthu K, Koh V, et al. SARS-CoV-2 seroprevalence and transmission risk factors among high-risk close contacts: a retrospective cohort study. *The Lancet Infectious Diseases*. 2021;21(3):333-343.

96.	Park SY, Kim Y-M, Yi S, et al. Coronavirus Disease Outbreak in Call Center, South Korea. *Emerging Infectious Diseases*. 2020;26(8)

97.	Park YJ, Choe YJ, Park O, et al. Contact Tracing during Coronavirus Disease Outbreak, South Korea, 2020. *Emerging Infectious Diseases*. October 2020;26(10)

98.	Peng J, Liu J, Mann SA, et al. Estimation of secondary household attack rates for emergent spike L452R SARS-CoV-2 variants detected by genomic surveillance at a community-based testing site in San Francisco. *Clinical Infectious Diseases*. 2021;doi:10.1093/cid/ciab283

99.	Pett J, McAleavey P, McGurnaghan P, et al. Epidemiology of COVID-19 in Northern Ireland, 26 February 2020–26 April 2020. *Epidemiology & Infection*. 2021;149

100.	Phiriyasart F, Chantutanon S, Salaeh F, et al. Outbreak Investigation of Coronavirus Disease (COVID-19) among Islamic Missionaries in Southern Thailand, April 2020. *OSIR Journal*. 2020;13(2)

101.	Reid MJA, Prado P, Brosnan H, et al. Assessing testing strategies and duration of quarantine in contact tracing for SARS-CoV-2: a retrospective study of San Francisco’s COVID-19 contact tracing program, June- August, 2020. *Open Forum Infectious Diseases*. 2021;doi:10.1093/ofid/ofab171

102.	Rosenberg ES, Dufort EM, Blog DS, et al. COVID-19 Testing, Epidemic Features, Hospital Outcomes, and Household Prevalence, New York State-March 2020. *Clin Infect Dis*. May 8 2020;doi:10.1093/cid/ciaa549

103.	Semakula M, Niragire F, Umutoni A, et al. The secondary transmission pattern of COVID-19 based on contact tracing in Rwanda. *BMJ Global Health*. 2021;6(6):e004885. doi:10.1136/bmjgh-2020-004885

104.	Seto J, Aoki Y, Komabayashi K, et al. Epidemiology of coronavirus disease 2019 in Yamagata Prefecture, Japan, January-May 2020: The importance of retrospective contact tracing. *Jpn J Infect Dis*. Mar 31 2021;doi:10.7883/yoken.JJID.2020.1073

105.	Son H, Lee H, Lee M, et al. Epidemiological characteristics of and containment measures for coronavirus disease 2019 in Busan Metropolitan City, South Korea. *Epidemiol Health*. Jun 1 2020:e2020035. doi:10.4178/epih.e2020035

106.	Shah K, Desai N, Saxena D, Mavalankar D, Mishra U, Patel GC. Household Secondary Attack Rate in Gandhinagar district of Gujarat state from Western India. *medRxiv*. 2020:2020.09.03.20187336. doi:10.1101/2020.09.03.20187336

107.	Sun WW, Ling F, Pan JR, et al. [Epidemiological characteristics of 2019 novel coronavirus family clustering in Zhejiang Province]. *Zhonghua Yu Fang Yi Xue Za Zhi*. Mar 15 2020;54(0):E027. doi:10.3760/cma.j.cn112150-20200227-00199

108.	Sunday V, Bhaskar E. Low secondary transmission rates of SARS-CoV-2 infection among contacts of construction laborers at open air environment. *Germs*. 2021;11(1):128-131.

109.	Tak P, Rohilla J. COVID-19 contact tracing in a tertiary care hospital: A retrospective chart review. *Infectious Disease Modelling*. 2021;6:1-4. doi:10.1016/j.idm.2020.10.014

110.	Telle K, Jorgensen SB, Hart RK, Greve-Isdahl M, Kacelnik O. Secondary attack rates of COVID-19 in Norwegian families: a nation-wide register-based study. *European Journal of Epidemiology*. 05/25 2021;doi:10.1007/s10654-021-00760-6

111.	Trunfio M, Longo BM, Alladio F, et al. On the SARS-CoV-2 “Variolation Hypothesis”: No Association Between Viral Load of Index Cases and COVID-19 Severity of Secondary Cases. Hypothesis and Theory. *Frontiers in Microbiology*. 2021-March-16 2021;12(473)doi:10.3389/fmicb.2021.646679

112.	Vallès X, Roure S, Valerio L, et al. SARS-CoV-2 contact tracing among disadvantaged populations during epidemic intervals should be a priority strategy: results from a pilot experiment in Barcelona. *Public Health*. 2021;

113.	van der Hoek W, Backer JA, Bodewes R, et al. [The role of children in the transmission of SARS-CoV-2]. *Nederlands tijdschrift voor geneeskunde*. Jun 3 2020;164De rol van kinderen in de transmissie van SARS-CoV-2.

114.	Verberk J, de Hoog M, Westerhof I, et al. Transmission of SARS-CoV-2 within households: a prospective cohort study in the Netherlands and Belgium – Interim results. *medRxiv*. 2021:2021.04.23.21255846. doi:10.1101/2021.04.23.21255846

115.	Wang X, Pan Y, Zhang D, et al. Basic epidemiological parameter values from data of real-world in mega-cities: the characteristics of COVID-19 in Beijing, China. *BMC Infectious Diseases*. 2020/07/20 2020;20(1):526. doi:10.1186/s12879-020-05251-9

116.	Wang Y, Tian H, Zhang L, et al. Reduction of secondary transmission of SARS-CoV-2 in households by face mask use, disinfection and social distancing: a cohort study in Beijing, China. *BMJ Global Health*. 2020;5(5):e002794.

117.	Wilkinson K, Chen X, Shaw S. Secondary attack rate of COVID-19 in household contacts in the Winnipeg Health Region, Canada. *Canadian Journal of Public Health*. 2021/02/01 2021;112(1):12-16. doi:10.17269/s41997-020-00451-x

118.	Wu J, Huang Y, Tu C, et al. Household Transmission of SARS-CoV-2, Zhuhai, China, 2020. *Clin Infect Dis*. May 11 2020;doi:10.1093/cid/ciaa557

119.	Wu P, Liu F, Chang Z, et al. Assessing asymptomatic, pre-symptomatic and symptomatic transmission risk of SARS-CoV-2. *Clinical Infectious Diseases*. 2021;doi:10.1093/cid/ciab271

120.	Wu Y, Song S, Kao Q, Kong Q, Sun Z, Wang B. Risk of SARS-CoV-2 infection among contacts of individuals with COVID-19 in Hangzhou, China. *Public Health*. 2020/08/01/ 2020;185:57-59. doi:<https://doi.org/10.1016/j.puhe.2020.05.016>

121.	Xin H, Jiang F, Xue A, et al. Risk factors associated with occurrence of COVID-19 among household persons exposed to patients with confirmed COVID-19 in Qingdao Municipal, China. *Transboundary and emerging diseases*. Jul 20 2020;doi:10.1111/tbed.13743

122.	Yung CF, Kam KQ, Chong CY, et al. Household Transmission of SARS-CoV-2 from Adults to Children. *The Journal of Pediatrics*. Jul 4 2020;doi:10.1016/j.jpeds.2020.07.009

123.	Zhang JZ, Zhou P, Han DB, et al. [Investigation on a cluster epidemic of COVID-19 in a supermarket in Liaocheng, Shandong province]. *Zhonghua Liu Xing Bing Xue Za Zhi*. Apr 27 2020;41(0):E055. doi:10.3760/cma.j.cn112338-20200228-00206

124.	Zhang W, Cheng W, Luo L, et al. Secondary Transmission of Coronavirus Disease from Presymptomatic Persons, China. *Emerging Infectious Diseases*. 2020;26(8)

125.	Zhuang YL, Zhang YT, Li M, et al. [Analysis on the cluster epidemic of coronavirus disease 2019 in Guangdong Province]. *Zhonghua Yu Fang Yi Xue Za Zhi*. Jul 6 2020;54(7):720-725. doi:10.3760/cma.j.cn112150-20200326-00446

126.	Tanaka ML, Marentes Ruiz CJ, Malhotra S, et al. SARS-CoV-2 Transmission Dynamics in Households With Children, Los Angeles, California. Original Research. *Frontiers in Pediatrics*. 2022-January-05 2022;9(1520)doi:10.3389/fped.2021.752993

127.	Lee M, Eun Y, Park K, Heo J, Son H. Follow-up investigation of asymptomatic COVID-19 cases at diagnosis in Busan, Korea. *Epidemiology and health*. 2020;42

128.	Arnedo-Pena A, Sabater-Vidal S, Meseguer-Ferrer N, et al. COVID-19 secondary attack rate and risk factors in household contacts in Castellon (Spain): Preliminary report. *Enfermedades Emergentes*. 2020;19(2):64-70.

129.	Carazo S, Laliberté D, Villeneuve J, et al. Characterization and evolution of infection control practices among SARS-CoV-2 infected healthcare workers of acute care hospitals and long-term care facilities in Quebec, Canada, Spring 2020. *Infection Control & Hospital Epidemiology*. 2021:1-37. doi:10.1017/ice.2021.160

130.	Patel A, Charani E, Ariyanayagam D, et al. New-onset anosmia and ageusia in adult patients diagnosed with SARS-CoV-2 infection. *Clin Microbiol Infect*. Jun 2 2020;doi:10.1016/j.cmi.2020.05.026

131.	Teherani MF, Kao CM, Camacho-Gonzalez A, et al. Burden of illness in households with SARS-CoV-2 infected children. *Journal of the Pediatric Infectious Diseases Society*. Aug 11 2020;doi:10.1093/jpids/piaa097

132.	Tibebu S, A. Brown K, Daneman N, Paul LA, Buchan SA. Household secondary attack rate of COVID-19 by household size and index case characteristics. *medRxiv*. 2021:2021.02.23.21252287. doi:10.1101/2021.02.23.21252287

133.	Fung HF, Martinez L, Alarid-Escudero F, et al. The household secondary attack rate of severe acute respiratory syndrome coronavirus 2 (SARS-CoV-2): a rapid review. *Clinical Infectious Diseases*. 2021;73(Supplement_2):S138-S145.
